# Consensus Risk Modeling and Uncertainty Quantification of Alzheimer’s Disease Using 5ADCSI Plasma Biomarkers and Multiple External Machine-Learning Frameworks

**DOI:** 10.64898/2026.07.30.26359341

**Authors:** Ebrahim Zandi, Sophie A. Bell, Eric Turkheimer, Deborah G. Finkel, Jonathan Becker, Deborah Winders Davis, Christopher R. Beam

**Affiliations:** University of Southern California, USA; Department of Immunology and Immune Therapeutics and Norris Cancer Comprehensive Cancer Center, Keck School of Medicine, Los Angeles, California 90033, USA; Department of Psychology, University of Virginia, Charlottesville, Virginia, 22904, USA; School of Health and Welfare, Institute of Gerontology, Jönköping University, Jönköping, Sweden; Department of Family and Geriatric Medicine, University of Louisville School of Medicine, Louisville, KY 40202 USA; Department of Pediatrics, University of Louisville School of Medicine, Louisville, KY 40202, USA; Department of Psychology, School of Gerontology, Los Angeles, California 90033, USA

**Keywords:** Alzheimer’s disease, plasma biomarkers, p217tau, 5ADCSI, machine learning, consensus risk, prediction uncertainty, amyloid PET, centiloids, APOE ε4, biomarker harmonization, assay platform effects, risk stratification, translational inference

## Abstract

**Background:** Blood-based biomarkers are increasingly used to identify Alzheimer’s disease (AD)-related pathology, but differences in p217tau assay methodology, training cohorts, and model-development context can substantially influence machine-learning (ML) predictions. Whether emerging biomarker platforms preserve biologically meaningful AD-related information across independently developed ML frameworks remains incompletely understood.

**Objective:** To evaluate the biological coherence and translational consistency of plasma biomarker measurements generated using the 5ADCSI platform by applying multiple externally trained ML frameworks and developing a consensus-risk approach that integrates framework predictions while quantifying prediction uncertainty.

**Methods:** Plasma biomarker measurements from 472 participants in the Louisville Twins Study were analyzed using three independently trained ML frameworks: an A4- derived model using the Lilly p217tau MSD assay and two ADNI-derived models using Quanterix Simoa p217tau measured with either the AlzPath or Janssen antibody.

Framework-specific predictions of amyloid positivity probability and predicted centiloid burden were integrated into consensus amyloid risk, consensus centiloid burden, and composite consensus AD-risk scores. Prediction uncertainty and rank instability were used to characterize framework agreement and participant-level classification stability.

**Results:** All three frameworks recognized biologically coherent AD-related signal despite differences in training cohort and assay methodology. Agreement was strongest between the A4-MSD and ADNI-AlzPath frameworks, whereas agreement involving the ADNI-Jan framework was weaker. Consensus-risk modeling identified a reproducibly high-risk subgroup characterized by elevated consensus-risk scores, low prediction uncertainty, and low rank instability. Participants prioritized by the consensus framework were enriched for APOE ε4 burden, p-tau217, p-tau217/Aβ42, and GFAP, while discordant high-risk participants exhibited substantially greater framework disagreement.

**Conclusions:** Plasma biomarker measurements generated using the 5ADCSI platform preserve biologically meaningful AD-related information that is consistently recognized across multiple independent ML frameworks. Consensus-risk modeling provides a practical strategy for integrating complementary information from external biological reference models while explicitly characterizing prediction uncertainty, thereby supporting evaluation of emerging blood-based biomarker platforms when direct pathological validation is unavailable.

## INTRODUCTION

The development of blood-based biomarkers has fundamentally transformed the landscape of Alzheimer’s disease (AD) research and clinical translation. For decades, the assessment of amyloid and tau pathology relied primarily on cerebrospinal fluid (CSF) measurements and positron emission tomography (PET) imaging, approaches that provide valuable biological information but are costly, invasive, and difficult to implement at scale. Recent advances in plasma biomarker technologies have created the possibility of inexpensive, minimally invasive screening tools capable of identifying individuals at elevated risk for AD-related pathology years before the onset of dementia symptoms^1–5^.

Among currently available plasma biomarkers, phosphorylated tau 217 (p217tau) has emerged as one of the most promising indicators of AD-related pathology.

Numerous studies have demonstrated strong associations between plasma p217tau concentrations and amyloid PET positivity, tau PET burden, CSF biomarkers, cognitive decline, and incident dementia. In many settings, p217tau-based blood tests approach the diagnostic performance of substantially more expensive imaging-based approaches and are increasingly being implemented in both clinical and research environments^4, 6–9^.

At the same time, the rapid expansion of blood-based biomarker technologies has highlighted an important translational challenge. Multiple p217tau assays are now available, including platforms that differ in antibody selection, analytical sensitivity, calibration procedures, dynamic range, and signal-processing methodology^7, 10–14^.

Although these assays target similar biological processes, measurable differences in biomarker distributions have been observed across platforms^7, 15, 16^. Such differences raise important questions regarding assay harmonization, comparability, and the portability of predictive models developed using one measurement system when applied to data generated using another.

These concerns extend beyond simple biomarker correlation. Increasingly, machine-learning (ML) approaches are being used to integrate plasma biomarkers, demographic variables, and genetic risk factors into multivariable prediction frameworks^17–21^. ML models have demonstrated strong performance for predicting amyloid PET positivity, continuous amyloid burden, disease stage, and future cognitive decline^22^. However, models optimized within one cohort or assay context may behave differently when transferred to new populations or measurement systems^21^. Differences in disease prevalence, cohort composition, biomarker distributions, and assay characteristics can influence prediction calibration, participant ranking, and downstream clinical interpretation.

Recently, we demonstrated that ML models trained in the Alzheimer’s Disease Neuroimaging Initiative (ADNI)^23^ and the Anti-Amyloid Treatment in Asymptomatic Alzheimer’s Disease (A4)^24^ cohorts using plasma biomarkers achieve strong performance for predicting amyloid PET positivity and continuous amyloid burden^21^. Importantly, those analyses showed that substantial biological signal remains detectable during pairwise external validation even when clinically actionable metrics are affected by dataset shift^21, 25^. Although model discrimination remained relatively stable across cohorts, calibration, participant prioritization, and negative predictive value were more sensitive to changes in cohort composition and prevalence structure^21^. These findings suggest that biologically meaningful disease-related information may persist across contexts even when quantitative agreement is incomplete.

This observation raises a broader question regarding evaluation of emerging biomarker platforms. Traditional validation strategies typically focus on direct assay correlation or prediction accuracy relative to a single reference standard^26^. However, such approaches provide limited insight into whether a novel measurement system preserves the broader biological structure recognized by independently developed disease models. An alternative strategy is to treat externally trained ML models as biological reference frameworks and evaluate whether a new biomarker platform generates coherent disease-related predictions across multiple independent models.

Importantly, biological plausibility and model agreement are not synonymous.

Different ML frameworks may recognize similar underlying disease-related biology while nevertheless producing different prediction scales, participant rankings, or risk estimates^27^. Conversely, strong agreement between two models does not necessarily imply biological validity. Therefore, understanding both prediction consistency and prediction uncertainty may be essential for translating emerging biomarker technologies into clinical practice^28^.

The 5ADCSI platform is a recently developed multiplex plasma biomarker assay that simultaneously quantifies multiple AD-related analytes, including p217tau, Aβ42, Aβ40, glial fibrillary acidic protein (GFAP), and neurofilament light chain (NfL)^29^. Initial analytical and clinical studies suggest that the platform captures biologically relevant AD-related information; however, its behavior when interpreted through independently trained ML frameworks is unknown.

In the present study, we applied three externally trained ML frameworks to cross-sectional plasma biomarker measurements generated using the 5ADCSI platform in an independent longitudinal aging cohort. One framework was trained in the A4 cohort using p217tau measured on the Lilly Meso Scale Discovery (MSD) platform^24, 30^. Two additional frameworks were trained in ADNI using p217tau measured on the Quanterix Simoa platform with either the AlzPath antibody^8^ or the Janssen antibody^31^. Rather than evaluating a single model, we used these frameworks as complementary biological reference lenses through which to interpret the same participant-level biomarker data.

The rationale for using multiple external frameworks was that if the biological information captured by the 5ADCSI platform is robust, independently developed ML frameworks should converge on similar disease-related biological patterns despite differences in training cohort and assay methodology.

Building on this approach, we developed a consensus-risk framework that integrates predictions across multiple external models while explicitly quantifying uncertainty arising from differences in training cohort, assay platform, and learned biomarker relationships. The framework was designed not only to estimate participant-level AD-related risk, but also to distinguish participants whose classification remained stable across models from those whose classification was highly framework dependent.

Specifically, we sought to determine whether 5ADCSI-derived plasma biomarker measurements generate biologically coherent predictions across independent machine-learning (ML) frameworks, whether a stable subgroup of participants with elevated AD-related biological risk can be consistently identified across models, whether disagreement among frameworks contains biologically meaningful information, and whether integrating predictions from multiple external frameworks provides a more informative characterization of AD-related biological risk than any individual model alone.

We hypothesized that if 5ADCSI measurements capture meaningful AD-related biological information, independently trained frameworks would converge on a common disease-related signal despite differences in training cohort and p217tau assay methodology. We further hypothesized that integrating predictions across multiple external reference frameworks would distinguish participants with stable AD-related biological risk from those whose classification remains sensitive to assay methodology or model-development context. More broadly, this work explores consensus machine-learning–based translational inference as a framework for evaluating emerging blood-based biomarker technologies and for developing future approaches to dementia-risk stratification when direct pathological validation is unavailable.

## METHODS

### Study Overview

Plasma biomarker measurements generated using the 5ADCSI platform were analyzed using three previously developed external machine-learning frameworks trained in the A4 and ADNI cohorts^21^. Framework-specific predictions of amyloid positivity probability and predicted centiloid burden were integrated into a consensus-risk framework that quantified participant-level AD-related biological risk, prediction uncertainty, and framework agreement. Subsequent analyses evaluated consensus-risk classification, participant prioritization, framework concordance, and biological correlates of consensus risk and framework disagreement.

## Study Cohorts

### Louisville Twins Study (LTS)

The Louisville Twins Study (LTS) is a longitudinal community-based twin and family cohort project established in 1958 and continuing through 1998 to investigate physiological, cognitive, and psychosocial development^32, 33^. Re-enrollment occurred in 2020-2024 with a new focus on lifespan cognitive development and risk factors associated with preclinical symptoms of Alzheimer’s disease and related dementias.

The present study included 472 LTS participants with available plasma Alzheimer’s disease biomarkers, APOE genotype, and demographic (i.e., sex, race/ethnicity, and education) data. Although the LTS is a longitudinal twin cohort, the present investigation did not examine twin-pair relationships or within-family effects. Instead, participants were analyzed as a single cohort because the objective of this study was to evaluate the biological coherence and translational consistency of 5ADCSI-derived plasma biomarker measurements across multiple externally trained machine-learning frameworks.

All study procedures were conducted in accordance with the Declaration of Helsinki and were approved by the Institutional Review Board at the University of Louisville (19.0989) and the University of Southern California (UP-20-00029). Written informed consent was obtained from all participants or their legally authorized representatives prior to participation.

### 5ADCSI Assay

Deidentified plasma samples were analyzed using the 5ADCSI multiplex blood biomarker platform as previously described by Alishahi et al. (2025)^34^. Plasma concentrations of phosphorylated tau 217(p217tau), amyloid-β42 (Aβ42), amyloid-β40 (Aβ40), glial fibrillary acidic protein (GFAP), and neurofilament light chain (NfL) were quantified in triplicate according to the validated 5ADCSI assay protocol^34^. APOE genotyping was performed using an in-house assay based on the method described by Zhong et al.^35^. Additional demographic variables included age, sex, race, and years of education.

Amyloid PET imaging was not available in the LTS cohort. Consequently, analyses focused on biological coherence, framework agreement, uncertainty quantification, and translational consistency rather than direct prediction accuracy.

### External Machine-Learning Reference Frameworks

Three previously developed machine-learning (ML) frameworks were used as external biological reference models^21^. The first framework was developed using data from the A4 study, a large prevention cohort composed primarily of cognitively unimpaired adults 65–85 years of age who underwent amyloid PET imaging as part of a preclinical Alzheimer’s disease prevention study^24^. The framework was developed using plasma p217tau measurements generated with the Lilly immunoassay on the MSD platform together with demographic and APOE variables^30^. The second framework was developed using data from ADNI, a longitudinal observational cohort^23^. ADNI includes older adults (55-90) across the Alzheimer’s disease continuum, including cognitively normal individuals, participants with mild cognitive impairment, and individuals with Alzheimer’s disease dementia, with extensive clinical, neuroimaging, genetic, and biomarker characterization. Data from plasma measured using the Quanterix Simoa platform with the AlzPath antibody (ADNI-AlzPath) was used for this study^8^. The third framework was also developed using ADNI data but utilized plasma measured on the Quanterix Simoa platform with the Janssen antibody (ADNI-Jan)^31^.

The three frameworks differed in both training-cohort composition and p-tau217 assay methodology, providing complementary external biological reference models for interpretation of the same 5ADCSI-derived biomarker measurements.

Each framework generated participant-level predictions of amyloid positivity probability and predicted amyloid burden expressed as centiloids. The ADNI-derived frameworks additionally generated diagnostic probability outputs for clinical disease stage. These exploratory outputs were used only in secondary analyses but were not incorporated into consensus-risk construction.

### Biomarker Data Preprocessing

Biomarker concentrations were log1p-transformed and z-score normalized during original machine-learning model development and prediction generation. For descriptive analyses and biological characterization, raw biomarker values were retained to preserve biological interpretability. Plasma biomarker concentrations were log-transformed for visualization only, and values are reported in original assay-specific units unless otherwise stated.

### Consensus-Risk Construction

Framework-specific prediction outputs were first standardized (z scores) to account for differences in numerical scale. Standardized amyloid-probability predictions were averaged to generate the consensus amyloid-risk score, and standardized predicted centiloid values were averaged to generate the consensus centiloid-burden score. The overall consensus AD-risk score was calculated as: Consensus AD-risk = (Consensus Amyloid Risk + Consensus Centiloid Burden) / 2. This approach was designed to identify AD-related biological signal that remained consistent across independently trained frameworks while reducing dependence on any single cohort or assay platform.

### Prediction Uncertainty and Rank Instability

Prediction uncertainty was quantified as the between-framework standard deviation of participant-level predictions. Amyloid uncertainty and centiloid uncertainty were calculated separately from framework-specific amyloid-probability and predicted-centiloid outputs, respectively. Total uncertainty was defined as: Total Uncertainty = (Amyloid Uncertainty + Centiloid Uncertainty) / 2.

Participants were ranked within each framework according to predicted amyloid probability and predicted centiloid burden. Rank instability was quantified as the standard deviation of participant rankings across frameworks, with lower values indicating more stable participant prioritization. Relationships among consensus AD- risk, prediction uncertainty, and rank instability were visualized to identify stable and unstable regions of the consensus-risk landscape (Figure 4).

### Consensus-Risk Group Classification

Participants were classified into four consensus-risk groups using consensus-risk magnitude and prediction uncertainty: Robust Low Risk, Intermediate, Robust High Risk, and Discordant High Risk. Robust High Risk participants exhibited high consensus risk with low prediction uncertainty, whereas Discordant High Risk participants exhibited high consensus risk with high prediction uncertainty.

### Consensus-Risk Ranking

Participants were ranked according to their composite consensus AD-risk score. The 20 highest-ranked participants were designated the Top 20 Consensus-Risk group for subsequent biological characterization (Figure 3).

### Biological Characterization of Highest-Risk Participants

The Top 20 Consensus-Risk participants were compared with the remainder of the cohort with respect to plasma p217tau, p217tau/Aβ42 ratio, Aβ42/Aβ40 ratio, GFAP, NfL, and APOE ε4 burden. Continuous variables were summarized as mean ± SD. Group differences were assessed using two-sided Mann-Whitney U tests because several biomarker distributions were non-normal. False-discovery-rate correction was performed using the Benjamini-Hochberg procedure across all variables included in the comparison.

### Biomarker and Covariate Profiles Across Consensus-Risk Groups

Biomarker and demographic characteristics were compared across the four consensus-risk groups (Robust Low Risk, Intermediate, Robust High Risk, and Discordant High Risk). Variables included p217tau, p217tau/Aβ42 ratio, Aβ42/Aβ40 ratio, GFAP, NfL, APOE ε4 burden, age, education, and sex.

### Framework Agreement Analyses

To evaluate the extent to which independently trained frameworks generated similar predictions when applied to identical LTS biomarker measurements, pairwise agreement analyses were performed between the three external reference frameworks. Agreement was evaluated separately for amyloid positivity probability and predicted centiloid burden. Pearson correlation coefficients quantified linear agreement, whereas Spearman correlations quantified rank-order agreement. Mean absolute error (MAE) and root mean squared error (RMSE) quantified numerical prediction differences between frameworks.

### Categorical Agreement Analyses

Predicted centiloid values were converted into categorical centiloid groups: Low (<20), Borderline (20–40), Elevated (40–80), High (>80). Agreement was quantified using both raw agreement percentages and Cohen’s κ statistics^36, 37^. These categorical analyses were exploratory and were not used for consensus-risk construction.

### High-Risk Overlap Analyses

Because clinical and research applications often prioritize identification of the highest-risk individuals rather than agreement across the full participant distribution, top- k overlap analyses were performed, an approach commonly used for evaluating concordance among ranked lists and prioritization systems^38^. Participants were ranked according to framework-specific amyloid probability and predicted centiloid outputs.

Overlap was calculated for the top 10 participants, top 25 participants, top 50 participants, and top 100 participants. Jaccard similarity and overlap percentages were calculated to quantify concordance of participant prioritization^39, 40^. Unlike correlation analyses, which evaluate agreement across the entire participant distribution, top-k overlap analyses determine whether independently trained frameworks converged on the same highest-risk participants^38^.

### Biomarker Contribution Analysis

To explore which biological variables were most strongly associated with framework-specific predictions, consensus AD-risk, prediction uncertainty, and rank instability, model-agnostic biomarker contribution analyses were performed using surrogate machine-learning models^41^. A surrogate model is an interpretable model trained to reproduce the predictions of an otherwise inaccessible prediction model. Because the original externally trained frameworks were not available for direct feature- importance extraction, surrogate random-forest regression models were trained to reconstruct framework-generated outputs using demographic variables, APOE ε4 burden, and plasma biomarker measurements from the LTS cohort^41^.

Separate surrogate models were trained for: framework-specific amyloid probability predictions, framework-specific predicted centiloid burden, consensus AD-risk, total uncertainty, and rank instability. Model fidelity was evaluated using five-fold cross-validation and summarized using the coefficient of determination (R²) and mean absolute error (MAE). Feature contributions were estimated using permutation importance. For each feature, values were randomly permuted while all other variables were held constant, and the resulting reduction in model performance was recorded.

Greater reductions in model performance indicated stronger association with the target prediction output^42^. Features evaluated included: p217tau, p217tau/Aβ42 ratio, Aβ42/Aβ40 ratio, APOE ε4 burden, age, sex, race, education, GFAP, and NfL. Contribution analyses were interpreted in two ways: (1) As biological correlates of stable consensus-risk estimates; and (2) As potential contributors to framework disagreement, prediction uncertainty, and rank instability.

Continuous variables are presented as mean ± standard deviation unless otherwise specified. Categorical variables are presented as counts and percentages. Group comparisons involving non-normally distributed variables were performed using two-sided Mann-Whitney U tests. False-discovery-rate correction was performed using the Benjamini-Hochberg procedure where appropriate. All analyses were performed using Python and standard scientific computing libraries.

## RESULTS

### Cohort Characteristics

The demographic, genetic, and plasma biomarker characteristics of the LTS cohort are summarized in Table 1. A total of 472 participants were included. The cohort had a mean age of 53.2 ± 12.0 years, mean educational attainment of 15.5 ± 2.3 years, and was predominantly female (57.4%) and White (99.4%). APOE genotype distribution was 71.6% ε4 non-carriers (0 alleles), 27.1% heterozygous carriers (1 allele), and 1.3% homozygous carriers (2 alleles), consistent with a community-based, middle-aged cohort rather than a dementia-enriched clinical sample.

**Table 1.**
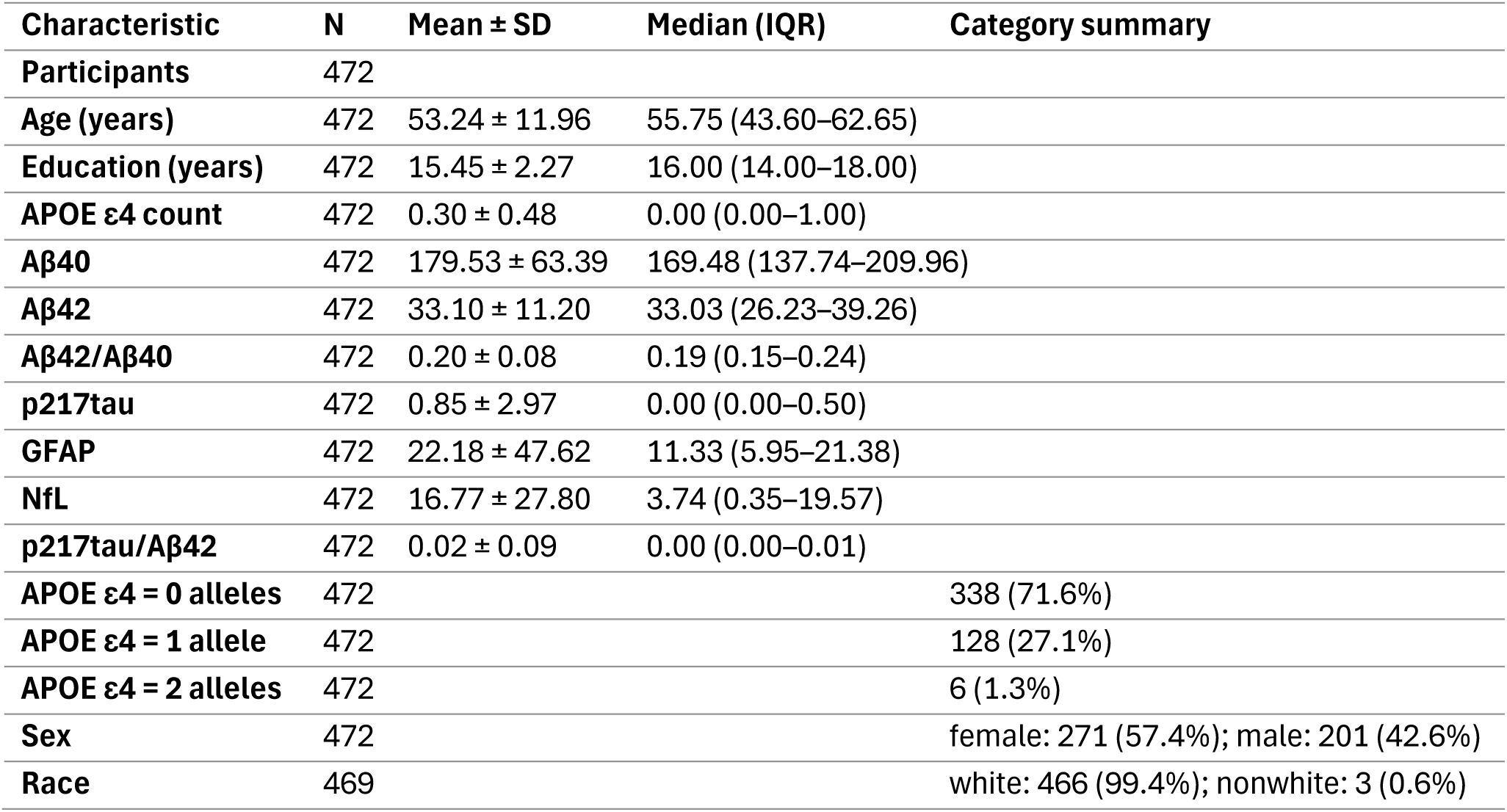
Characteristics of the Longitudinal Twins Study (LTS) Cohort. Demographic, genetic, and plasma biomarker characteristics of participants included in the Longitudinal Twins Study (LTS) cohort. Continuous variables are presented as mean ± standard deviation (SD) and median (interquartile range [IQR]). Categorical variables are presented as counts and percentages. Age was calculated from date of birth and expressed in years. Education was converted from the original coded educational categories to years of education. APOE genotype is summarized as the percentage of participants carrying 0, 1, or 2 ε4 alleles. Plasma biomarker measurements were obtained using the 5ADCSI platform and included Aβ40, Aβ42, p217tau, glial fibrillary acidic protein (GFAP), neurofilament light chain (NfL), Aβ42/Aβ40 ratio, and p217tau/Aβ42 ratio. This table provides a descriptive overview of the study population and establishes the biological and demographic context for the consensus-risk analyses. Because all subsequent machine-learning predictions were generated from these biomarker measurements, this table represents the baseline characteristics of the cohort rather than model-derived outputs.

| Characteristic | N | Mean ± SD | Median (IQR) | Category summary |
| --- | --- | --- | --- | --- |
| **Participants** | 472 |  |  |  |
| **Age (years)** | 472 | 53.24 ± 11.96 | 55.75 (43.60–62.65) | |
| **Education (years)** | 472 | 15.45 ± 2.27 | 16.00 (14.00–18.00) | |
| **APOE ε4 count** | 472 | 0.30 ± 0.48 | 0.00 (0.00–1.00) | |
| **Aβ40** | 472 | 179.53 ± 63.39 | 169.48 (137.74–209.96) | |
| **Aβ42** | 472 | 33.10 ± 11.20 | 33.03 (26.23–39.26) | |
| **Aβ42/Aβ40** | 472 | 0.20 ± 0.08 | 0.19 (0.15–0.24) | |
| **p217tau** | 472 | 0.85 ± 2.97 | 0.00 (0.00–0.50) | |
| **GFAP** | 472 | 22.18 ± 47.62 | 11.33 (5.95–21.38) | |
| **NfL** | 472 | 16.77 ± 27.80 | 3.74 (0.35–19.57) | |
| **p217tau/Aβ42** | 472 | 0.02 ± 0.09 | 0.00 (0.00–0.01) | |
| **APOE ε4 = 0 alleles** | 472 |  |  | 338 (71.6%) |
| **APOE ε4 = 1 allele** | 472 |  |  | 128 (27.1%) |
| **APOE ε4 = 2 alleles** | 472 |  |  | 6 (1.3%) |
| **Sex** | 472 |  |  | female: 271 (57.4%); male: 201 (42.6%) |
| **Race** | 469 |  |  | white: 466 (99.4%); nonwhite: 3 (0.6%) |

Plasma biomarker measurements obtained using the 5ADCSI platform showed substantial interindividual variability. Mean Aβ40 concentration was 179.5 ± 63.4, mean Aβ42 concentration was 33.1 ± 11.2, and the mean Aβ42/Aβ40 ratio was 0.20 ± 0.08. Mean p217tau concentration was 0.85 ± 2.97, remaining low overall as expected for a relatively healthy community-based cohort but demonstrating broad variability across participants. The mean p217tau/Aβ42 ratio was 0.02 ± 0.09, likewise showing considerable interindividual variation. Mean GFAP concentration was 22.2 ± 47.6, and mean NfL concentration was 16.8 ± 27.8, indicating a wide biological range in astroglial and neuroaxonal injury markers.

### Construction of a Consensus AD-Risk Landscape

Framework-specific predictions from the three external machine-learning models were first summarized to characterize their native prediction distributions (Supplementary Table S1) and subsequently integrated into a composite consensus AD-risk landscape (Figure 1). Because the three frameworks generated prediction outputs on different numerical scales (Supplementary Table S1), framework-specific outputs were standardized before constructing the consensus amyloid-risk and consensus centiloid-burden scores. The resulting consensus-risk landscape provides a participant-level representation of AD-related biological risk across the LTS cohort.

**Figure 1.**
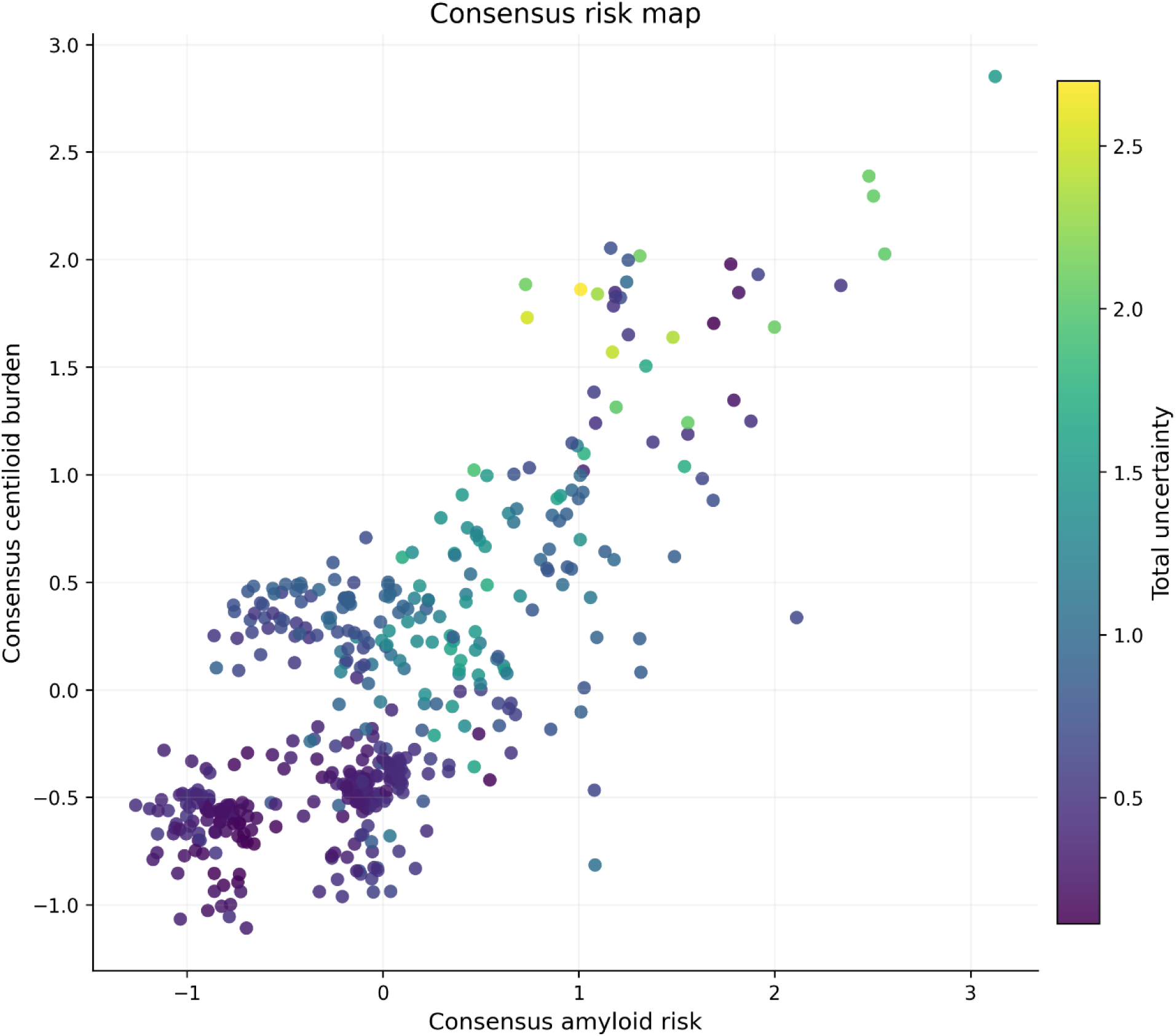
Consensus AD-Risk Map. Participant-level visualization of the consensus-risk framework derived from three independently trained machine-learning reference models. The x-axis shows consensus amyloid risk and the y-axis shows consensus centiloid burden. Both scores were generated by integrating framework-specific predictions from an A4-trained model using MSD p217tau, an ADNI-trained model using AlzPath p217tau, and an ADNI-trained model using Jan p217tau. Each point represents one participant in the LTS cohort. Point color indicates total prediction uncertainty, calculated from disagreement among framework-specific amyloid probability and centiloid predictions. Warmer colors indicate greater framework disagreement and therefore lower confidence in the resulting risk estimate. Participants occupying the upper-right region of the plot exhibit both elevated consensus amyloid risk and elevated consensus centiloid burden and therefore represent the biologically highest-risk portion of the cohort. In contrast, participants located in the lower-left region demonstrate low consensus risk across both dimensions. The concentration of uncertainty within intermediate regions of the risk landscape illustrates that biological risk and prediction confidence are distinct but complementary dimensions of interpretation. This figure provides the conceptual foundation for the consensus-risk framework by simultaneously visualizing risk magnitude and uncertainty.

The consensus-risk landscape demonstrated a strong positive relationship between consensus amyloid risk and consensus centiloid burden (Figure 1).

Participants occupying the upper-right region exhibited the highest consensus AD-risk, whereas those in the lower-left region showed consistently low estimated risk. The continuous gradient between these extremes indicates that consensus risk spans a broad spectrum of AD-related biological variation rather than discrete high- and low-risk states.

Prediction uncertainty was lowest among participants with consistently low or high consensus-risk scores and highest among participants occupying intermediate regions of the landscape, indicating greater disagreement among the external frameworks in these regions.

Consensus AD-risk scores were positively skewed (Supplementary Figure S1), with most participants clustering at low-to-intermediate risk and a smaller subset extending into a high-risk tail.

### Consensus Risk Classification Identified Distinct Biological and Stability Profiles

Participants were classified into four consensus-risk groups based on consensus AD-risk and prediction uncertainty: Robust Low Risk, Intermediate, Robust High Risk, and Discordant High Risk (Figure 2; Table 2; Supplementary Table S2).

**Figure 2.**
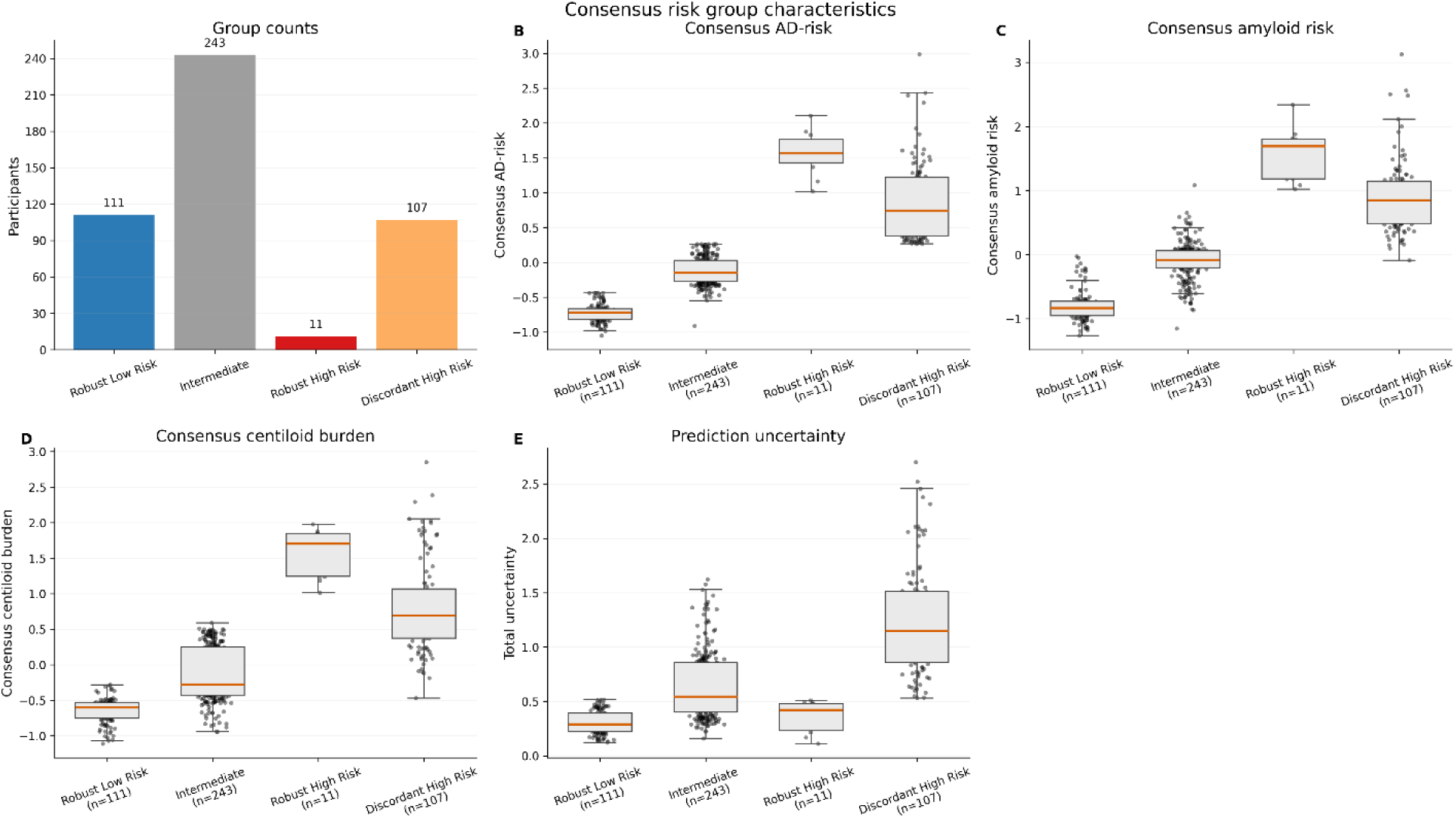
Characteristics of Consensus AD-Risk Groups. Multi-panel summary of the biological and statistical properties of the four consensus- risk groups: Robust Low Risk, Intermediate, Robust High Risk, and Discordant High Risk. (A) Number of participants assigned to each consensus-risk group. (B) Distribution of composite consensus AD-risk scores. (C) Distribution of consensus amyloid-risk scores. (D) Distribution of consensus centiloid-burden scores. (E) Distribution of total prediction uncertainty. Consensus AD-risk was calculated as the average of consensus amyloid risk and consensus centiloid burden. Consensus amyloid risk was derived from standardized framework-specific amyloid probability predictions, whereas consensus centiloid burden was derived from standardized framework-specific predicted centiloid values. Total uncertainty was calculated as the average of amyloid uncertainty and centiloid uncertainty and reflects disagreement among the three external machine- learning frameworks. The figure demonstrates that risk magnitude and prediction stability are distinct dimensions of interpretation. Robust High Risk participants exhibit elevated consensus risk together with low uncertainty, whereas Discordant High Risk participants exhibit similarly elevated risk but substantially greater framework disagreement. Consequently, the consensus-risk framework separates biologically stable high-risk participants from those whose classification remains framework dependent.

**Figure 3.**
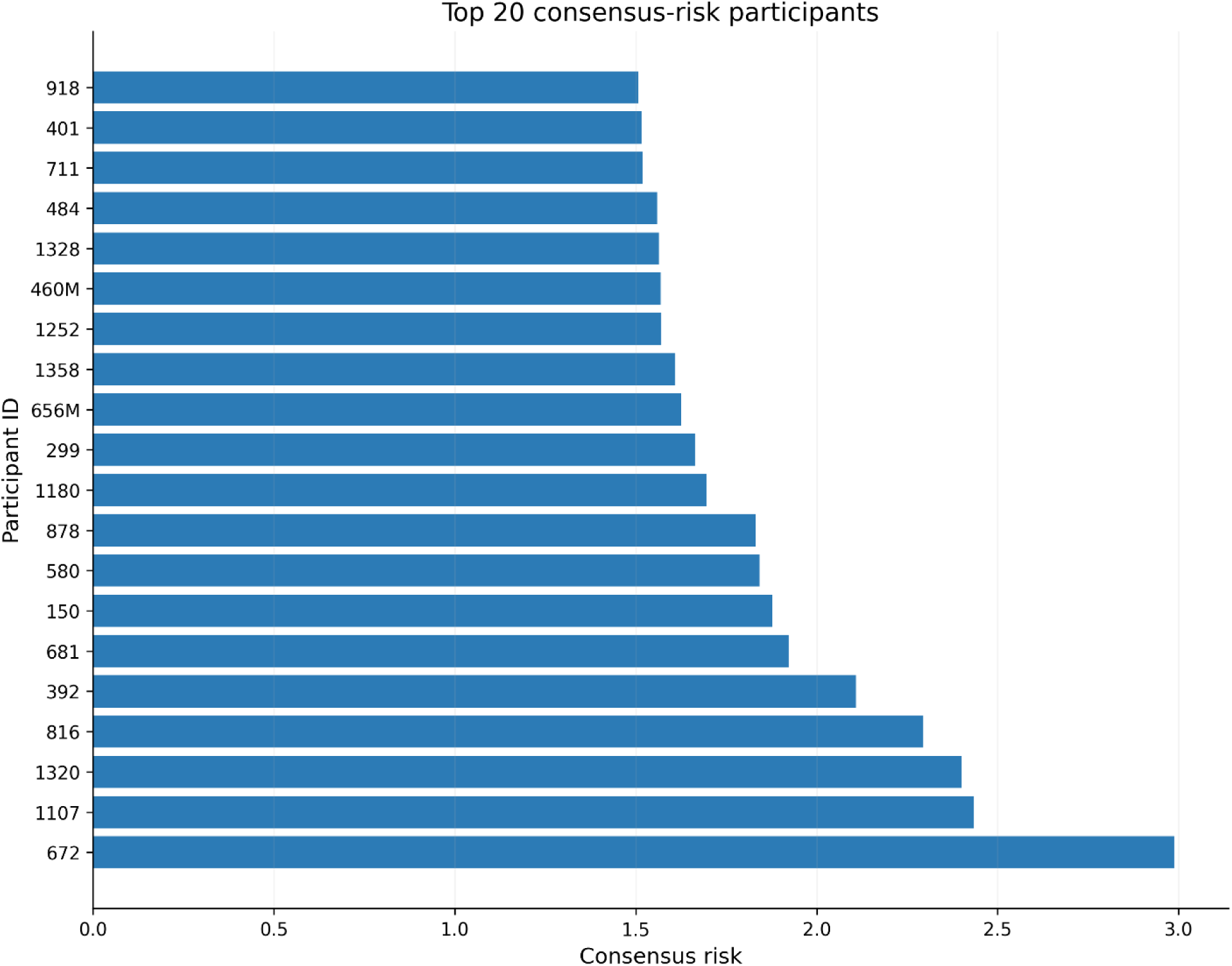
Top 20 Participants Ranked by Consensus AD-Risk. The twenty participants with the highest composite consensus AD-risk scores in the LTS cohort. Consensus AD-risk was generated by integrating framework-specific predictions from the A4-MSD, ADNI-AlzPath, and ADNI-Jan machine-learning models. Higher values indicate participants whose plasma biomarker profiles are most consistently associated with elevated AD-related biological risk across multiple external reference frameworks. These participants represent the highest-ranked subgroup identified by the consensus-risk framework and therefore constitute the most biologically enriched participants according to the integrated machine-learning approach. Unlike framework- specific rankings, which may be influenced by training cohort composition or assay platform, the consensus ranking reflects AD-related biological signal that remains elevated across multiple independent models.

**Figure 4.**
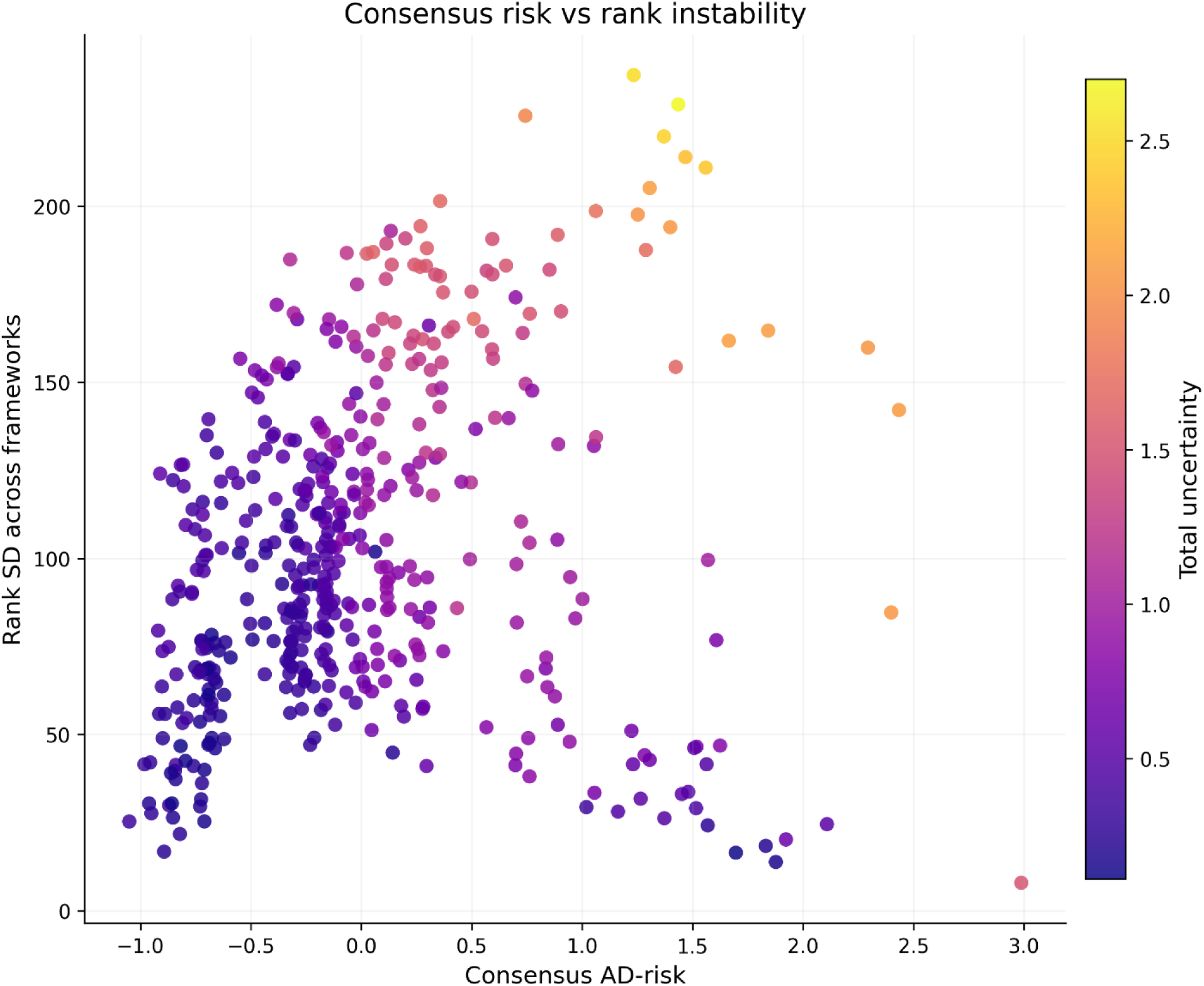
Consensus AD-Risk, Rank Instability, and Prediction Uncertainty. Relationship between consensus AD-risk, rank instability, and prediction uncertainty across the three external machine-learning frameworks. Each point represents one participant in the LTS cohort. The x-axis shows the composite consensus AD-risk score. The y-axis shows rank instability, quantified as the standard deviation of participant rankings across frameworks. Point color represents total prediction uncertainty. Participants with low rank instability receive similar rankings regardless of which machine-learning framework is used, whereas participants with high rank instability exhibit framework-dependent prioritization. The figure demonstrates that elevated biological risk does not necessarily imply high uncertainty. Some participants exhibit both high consensus risk and strong ranking stability, whereas others demonstrate substantial disagreement among frameworks despite similar average risk estimates. These findings support the use of uncertainty measures as an additional dimension of interpretation beyond risk magnitude alone.

**Table 2.** Characteristics of Consensus AD-Risk Groups. Comparison of participant characteristics across the four consensus AD-risk groups derived from the consensus machine-learning framework: Robust Low Risk, Intermediate, Robust High Risk, and Discordant High Risk. Participants were classified according to two dimensions: (1) the magnitude of their composite consensus AD-risk score and (2) the degree of agreement among machine-learning frameworks. Consensus AD-risk was calculated as the average of consensus amyloid risk and consensus centiloid burden. Consensus amyloid risk was generated by averaging standardized amyloid probability predictions from the A4-MSD, ADNI-AlzPath, and ADNI-Jan frameworks. Consensus centiloid burden was generated by averaging standardized predicted centiloid values across the same frameworks. Total uncertainty was calculated as the average of amyloid uncertainty and centiloid uncertainty, where uncertainty was defined as the standard deviation of framework-specific predictions. Rank instability was quantified using the standard deviation of participant rankings across frameworks. Values are presented as mean ± standard deviation (SD). Percentages indicate the proportion of the entire cohort represented by each risk group.

| Risk Group | N | Cohort (%) | Consensus AD-risk | Consensus amyloid risk | Consensus centiloid burden | Total uncertainty | Rank SD |
| --- | --- | --- | --- | --- | --- | --- | --- |
| Robust Low Risk | 111 | 23.50% | -0.72 ± 0.13 | -0.80 ± 0.25 | -0.64 ± 0.17 | 0.31 ± 0.11 | 74.48 ± 30.99 |
| Intermediate | 243 | 51.50% | -0.12 ± 0.20 | -0.10 ± 0.29 | -0.14 ± 0.39 | 0.64 ± 0.31 | 107.90 ± 34.91 |
| Robust High Risk | 11 | 2.30% | 1.56 ± 0.31 | 1.57 ± 0.41 | 1.55 ± 0.35 | 0.36 ± 0.15 | 25.95 ± 7.94 |
| Discordant High Risk | 107 | 22.70% | 0.86 ± 0.54 | 0.90 ± 0.55 | 0.81 ± 0.65 | 1.24 ± 0.50 | 125.57 ± 58.42 |

The Intermediate group was the largest category (n = 243; 51.5% of the cohort), followed by Robust Low Risk (n = 111; 23.5%) and Discordant High Risk (n = 107; 22.7%). Only 11 participants (2.3%) met criteria for Robust High Risk, indicating that strong agreement across all three external frameworks occurred in a relatively small subset of individuals. Consensus AD-risk measures clearly separated the four groups, with Robust Low Risk participants exhibiting the lowest consensus-risk scores and Robust High Risk participants exhibiting the highest scores (Figure 2B–D; Table 2).

The component scores underlying the composite consensus-risk framework followed a similar pattern. Consensus AD-risk increased from −0.74 ± 0.13 in the Robust Low Risk group to 1.61 ± 0.32 in the Robust High Risk group. Consensus amyloid risk increased from −0.84 ± 0.22 to 1.58 ± 0.42, while consensus centiloid burden increased from −0.63 ± 0.16 to 1.64 ± 0.34. Discordant High Risk participants also demonstrated elevated consensus amyloid risk and centiloid burden, indicating that they occupied the biologically high-risk portion of the risk landscape despite substantial disagreement among frameworks regarding the magnitude of risk.

Robust Low Risk and Robust High Risk participants both exhibited relatively low total uncertainty (0.31 ± 0.11 and 0.36 ± 0.15, respectively), whereas Discordant High Risk participants demonstrated more than threefold higher uncertainty (1.24 ± 0.50), indicating substantially greater disagreement among the external frameworks (Figure 2D; Table 2). Detailed analyses of participant-level rank instability across the consensus-risk landscape are presented in Figure 4.

### Biological Validation of the Highest Consensus-Risk Participants

The 20 participants with the highest consensus AD-risk scores exhibited substantially greater consensus AD-risk, consensus amyloid risk, and consensus centiloid burden than the remainder of the cohort (Figure 3; Table 3). As expected, all three consensus- derived measures differed markedly between groups (all FDR-adjusted q < 0.001), confirming that the consensus framework consistently prioritized participants with elevated predicted AD-related biological risk.

**Table 3.** Characteristics of the Highest Consensus-Risk Participants Compared With the Remaining Cohort. Comparison of demographic, genetic, biomarker, and consensus-risk characteristics between the 20 participants with the highest composite consensus AD-risk scores and the remainder of the LTS cohort. Participants were ranked according to their composite consensus AD-risk score, and the twenty highest-ranked individuals were designated the Top 20 Consensus-Risk group. The Remaining Cohort consisted of all other participants. Consensus AD-risk was calculated as the average of consensus amyloid risk and consensus centiloid burden. Consensus amyloid risk and consensus centiloid burden were generated from standardized framework-specific predictions derived from the A4-MSD, ADNI-AlzPath, and ADNI-Jan models. Values are reported as mean ± standard deviation (SD). Group comparisons were performed using two-sided Mann—Whitney U tests because several biomarker distributions were non-normal. False-discovery-rate (FDR) correction was applied using the Benjamini-Hochberg procedure across all variables included in the table. Columns are defined as follows: Top 20 Mean ± SD. Mean and SD for participants with the highest consensus-risk scores. Remaining Mean ± SD. Mean and SD for all remaining participants. P value: Mann—Whitney U test comparing the two groups. FDR q value: Benjamini—Hochberg adjusted q value controlling for multiple comparisons.

| Variable | Top 20 N | Top 20 Mean ± SD | Remaining N | Remaining Mean ± SD | P value | FDR q value |
| --- | --- | --- | --- | --- | --- | --- |
| Consensus AD-risk | 20 | 1.85 ± 0.40 | 452 | -0.08 ± 0.55 | <0.001 | <0.001 |
| Consensus amyloid risk | 20 | 1.80 ± 0.57 | 452 | -0.08 ± 0.62 | <0.001 | <0.001 |
| Consensus centiloid burden | 20 | 1.91 ± 0.35 | 452 | -0.08 ± 0.59 | <0.001 | <0.001 |
| Total uncertainty | 20 | 1.04 ± 0.79 | 452 | 0.68 ± 0.44 | 0.097 | 0.129 |
| Age (years) | 20 | 53.27 ± 10.14 | 452 | 53.23 ± 12.05 | 0.941 | 0.941 |
| Education (years) | 20 | 14.45 ± 2.19 | 452 | 15.50 ± 2.27 | 0.043 | 0.065 |
| APOE ε4 count | 20 | 0.80 ± 0.62 | 452 | 0.27 ± 0.47 | <0.001 | <0.001 |
| p217tau | 20 | 8.71 ± 10.49 | 452 | 0.50 ± 1.31 | <0.001 | <0.001 |
| Aβ42/Aβ40 | 20 | 0.20 ± 0.05 | 452 | 0.20 ± 0.08 | 0.416 | 0.499 |
| GFAP | 20 | 139.26 ± 172.99 | 452 | 17.00 ± 21.75 | <0.001 | <0.001 |
| NfL | 20 | 17.19 ± 21.44 | 452 | 16.76 ± 28.06 | 0.776 | 0.847 |
| p217tau/Aβ42 | 20 | 0.26 ± 0.34 | 452 | 0.01 ± 0.04 | <0.001 | <0.001 |

The Top 20 participants were also strongly enriched for established AD-related biological markers. APOE ε4 burden was nearly threefold higher than in the remainder of the cohort (0.80 ± 0.62 vs. 0.27 ± 0.47; q < 0.001). Mean plasma p217tau concentration was approximately 17-fold higher (8.71 ± 10.49 vs. 0.50 ± 1.31; q < 0.001), and the p217tau/Aβ42 ratio was more than 25-fold higher (0.26 ± 0.34 vs. 0.01 ± 0.04; q < 0.001). GFAP concentrations were likewise markedly elevated (139.26 ± 172.99 vs. 17.00 ± 21.75; q < 0.001), indicating enrichment for astroglial activation.

Together, these findings demonstrate that participants prioritized by the consensus framework were enriched for multiple independent biological markers associated with Alzheimer’s disease.

In contrast, several variables showed little or no evidence of enrichment. Mean age was essentially identical between groups (53.27 ± 10.14 vs. 53.23 ± 12.05 years; q = 0.941), indicating that the consensus framework was not simply selecting older individuals within this middle-aged cohort. Neither the Aβ42/Aβ40 ratio nor NfL differed significantly between groups after multiple-comparison correction, and although education was modestly lower among the Top 20 participants, this difference did not remain significant after false-discovery-rate correction (q = 0.065). Total prediction uncertainty was numerically higher among the Top 20 participants but likewise did not differ significantly after correction (q = 0.129). The complete participant-level consensus-risk rankings, including consensus risk, uncertainty, rank instability, and risk-group assignment for all participants, are provided in Supplementary Table S3.

### Framework Agreement Reflects Both Assay and Cohort Differences

To determine whether independently trained frameworks produced comparable predictions when applied to identical LTS biomarker measurements, pairwise agreement analyses were performed across all framework combinations (Supplementary Tables S4 and Figure S4). Because the frameworks differed with respect to both training cohort and assay methodology, these comparisons provide insight into the relative influence of biological signal, cohort composition, and assay platform. The strongest agreement was observed between the A4-MSD and ADNI- AlzPath frameworks (Supplementary Tables S4). Amyloid probability predictions demonstrated moderate agreement (Pearson r = 0.61), whereas predicted centiloid burden showed stronger agreement (Pearson r = 0.73). Despite being developed using different cohorts and different assay platforms, these two frameworks frequently produced similar participant rankings and identified many of the same individuals as high risk.

Agreement involving the ADNI-Jan framework was substantially weaker.

Comparisons between A4-MSD and ADNI-Jan demonstrated little agreement for either amyloid probability (r = 0.01) or predicted centiloid burden (r = −0.05) (Supplementary Tables S4). Agreement between ADNI-AlzPath and ADNI-Jan was modest for amyloid probability (r = 0.24) and low for predicted centiloid burden (r = 0.05). Importantly, the ADNI-AlzPath versus ADNI-Jan comparison largely isolates the effect of antibody selection because both models were trained within ADNI using the same Quanterix Simoa platform. The persistence of weaker agreement in this comparison suggests that antibody-specific differences in measurement can materially influence downstream machine-learning predictions.

### Categorical Agreement Was Highly Sensitive to Threshold Effects

To complement the continuous prediction analyses, we evaluated agreement for categorical centiloid classifications derived from framework-specific predicted centiloid values (Supplementary Table S5). This analysis was intended to determine whether frameworks that showed agreement in continuous predictions would also assign participants to the same clinically interpretable centiloid category after thresholding.

Despite moderate agreement for continuous predictions, categorical agreement was uniformly poor across framework pairs (Supplementary Table S5). The results differed substantially from the continuous prediction analyses. Agreement between the A4-MSD and ADNI-AlzPath frameworks was low (3.0%), while agreement between A4- MSD and ADNI-Jan was 10.8%. In contrast, agreement between the two ADNI-based frameworks reached 62.5%. However, despite these differences in raw agreement, Cohen’s κ values were close to zero for all framework pairs (κ range: −0.052 to 0.033), indicating that much of the apparent agreement was attributable to category prevalence rather than true participant-level concordance.

These findings contrast sharply with the continuous prediction analyses, in which moderate correlations and substantial high-risk overlap were observed between some framework pairs. The discrepancy suggests that converting continuous prediction outputs into discrete centiloid categories results in substantial information loss and can obscure biologically meaningful similarities between frameworks. Consequently, participants with similar underlying risk estimates may be assigned to different categories simply because their predicted values fall on opposite sides of an arbitrary threshold. Taken together, these results support the use of continuous prediction outputs for consensus-risk construction and indicate that category-based agreement metrics provide a less reliable representation of framework concordance than continuous prediction or participant-ranking approaches.

### Rank Instability and Prediction Uncertainty Varied Across the Consensus-Risk Landscape

To examine where framework disagreement was concentrated within the consensus-risk space, we plotted composite consensus AD-risk against rank instability, with points colored according to total prediction uncertainty (Figure 4). This analysis was designed to determine whether participant prioritization remained stable across frameworks throughout the risk spectrum or whether disagreement was concentrated within specific regions of the consensus-risk landscape.

Participants with low consensus-risk scores generally exhibited relatively low rank instability and low uncertainty, indicating strong agreement across frameworks regarding their biological classification (Figure 4). In contrast, participants occupying intermediate-risk regions demonstrated substantially greater rank instability and uncertainty, suggesting that framework-specific assumptions had a larger influence on participant ranking within this portion of the risk distribution.

The highest-risk region of the landscape was more heterogeneous. Some participants with elevated consensus-risk scores exhibited low rank instability and low uncertainty, indicating that they were consistently prioritized across frameworks. Other participants with similarly high consensus-risk values demonstrated substantially greater instability and uncertainty, reflecting marked disagreement among frameworks despite broadly similar average risk estimates. This pattern is consistent with the distinction between Robust High Risk and Discordant High Risk participants identified in Figure 2.

Overall, these findings indicate that uncertainty is not uniformly distributed across the risk spectrum. Rather than reflecting random prediction noise, framework disagreement appears concentrated among participants whose biomarker profiles occupy biologically ambiguous or model-sensitive regions of the consensus-risk landscape. The coexistence of stable and unstable high-risk participants further supports the value of incorporating uncertainty measures alongside consensus-risk estimates when prioritizing individuals for follow-up evaluation.

### Identification of High-Risk Participants Revealed Substantial Differences Across Frameworks

Top-k overlap analyses revealed substantial differences in participant prioritization across the three frameworks (Figure 5; Table S6). The strongest overlap among high-risk participants was observed between the A4-MSD and ADNI-AlzPath frameworks. For amyloid probability predictions, overlap ranged from 60% among the top 10 participants to 80% among the top 25 participants. Similar patterns were observed for predicted centiloid burden, where overlap reached 83% among the top 100 participants. These findings indicate that the two frameworks frequently identified the same participants as highest risk despite being developed in different cohorts and using different assay platforms.

**Figure 5.**
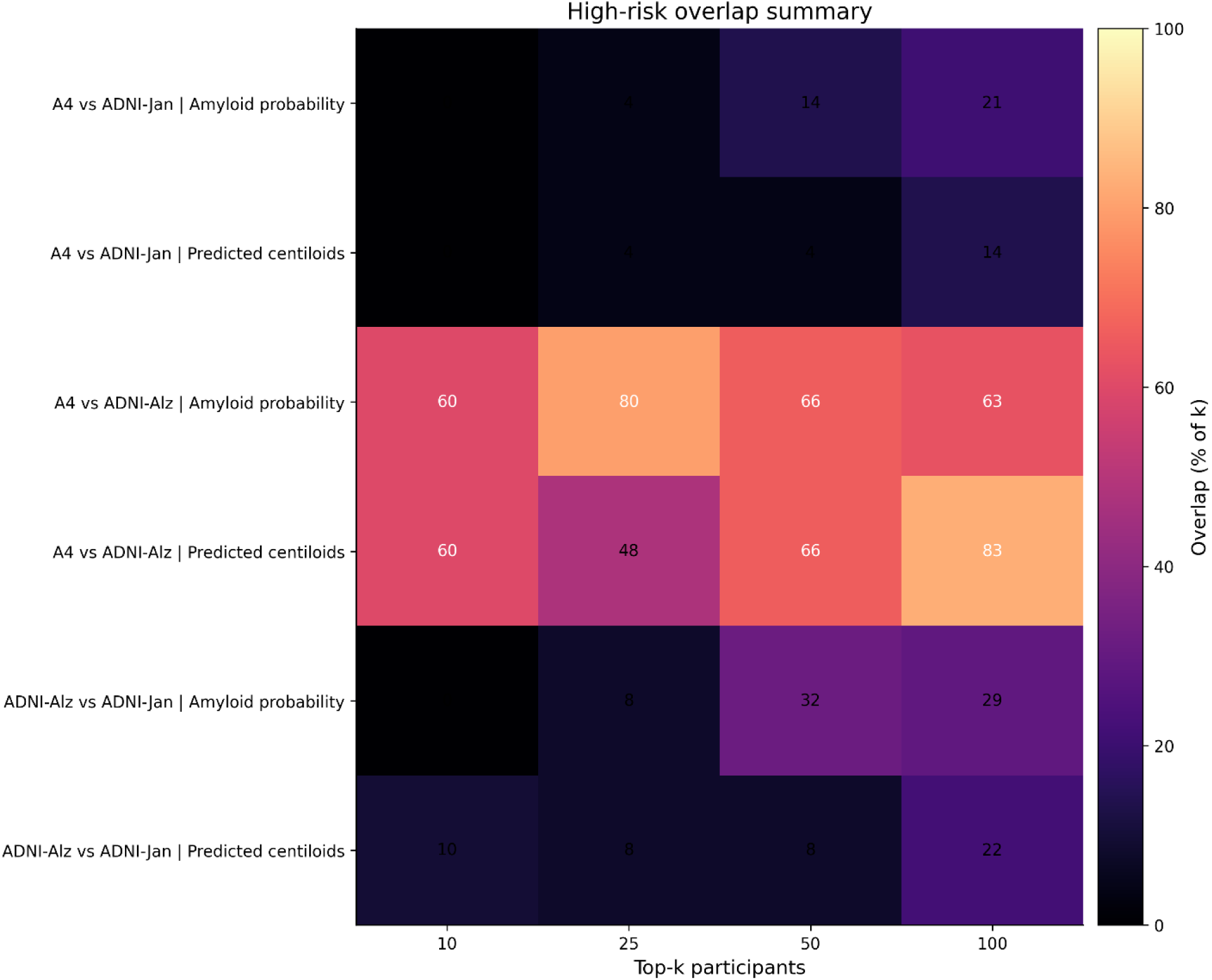
High-Risk Participant Overlap Across External Reference Frameworks. Heatmap summarizing overlap among participants identified as highest risk by the three external machine-learning frameworks. Rows represent pairwise framework comparisons for amyloid probability predictions and predicted centiloid burden. Columns correspond to increasingly inclusive definitions of the highest-risk subgroup (top 10, 25, 50, and 100 ranked participants). Values within each cell represent the percentage of participants shared between frameworks relative to the selected threshold. The strongest overlap is observed between the A4-MSD and ADNI-AlzPath frameworks, indicating substantial agreement regarding participant prioritization. In contrast, overlap involving the ADNI-Jan framework is considerably weaker, suggesting greater divergence in participant ranking. Because all frameworks were applied to identical LTS biomarker measurements, these findings demonstrate that training cohort composition and p217tau assay platform influence identification of high-risk individuals. The overlap analysis provides a clinically relevant measure of agreement because it directly evaluates whether different frameworks prioritize the same participants for potential enrichment, monitoring, or follow-up studies.

In contrast, overlap involving the ADNI-Jan framework was substantially lower.

Comparisons between A4-MSD and ADNI-Jan demonstrated minimal overlap across all ranking thresholds, while overlap between ADNI-AlzPath and ADNI-Jan remained modest despite both frameworks being trained within ADNI and using the same Quanterix Simoa platform. Thus, frameworks sharing the same cohort and platform did not necessarily identify the same high-risk participants.

These results suggest that differences in measurement methodology, including antibody selection, may substantially influence downstream participant prioritization. The consensus-risk framework addresses this challenge by identifying participants whose elevated risk remains robust across multiple independently trained models rather than relying on a single framework-specific ranking.

### Biomarker and Covariate Profiles Across Consensus Risk Groups

To determine whether the consensus-risk groups differed in biologically meaningful ways beyond the model outputs themselves, we compared plasma biomarker and covariate profiles across the four consensus-risk groups (Figure 6). This analysis was designed to evaluate whether the consensus framework identified participant subgroups that corresponded to known AD-related biological characteristics.

**Figure 6.**
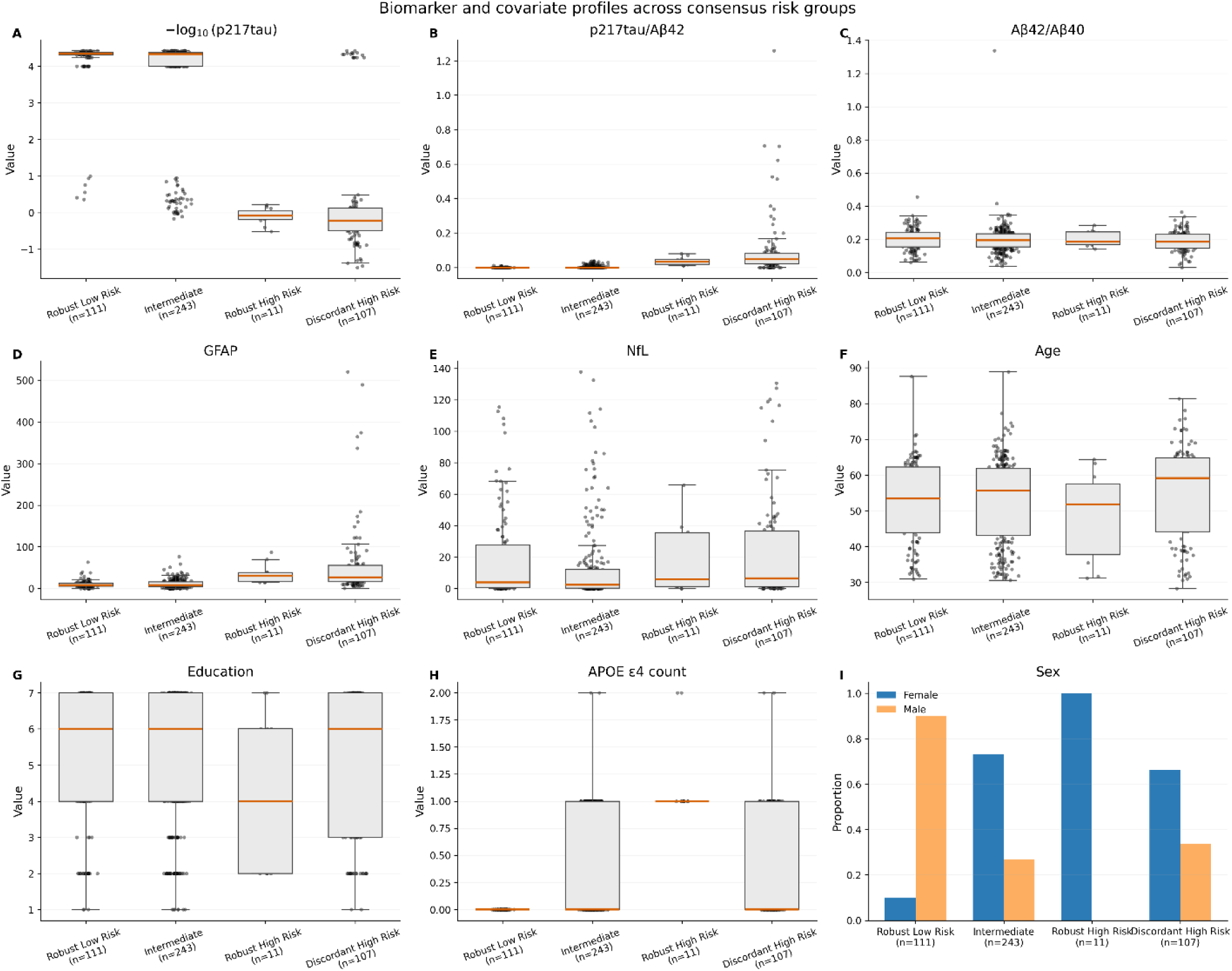
Biomarker and covariate profiles across consensus AD-risk groups. Distribution of plasma biomarkers and participant characteristics across the four consensus AD-risk groups: Robust Low Risk (n = 111), Intermediate (n = 243), Robust High Risk (n = 11), and Discordant High Risk (n = 107). Boxplots display the median (horizontal line), interquartile range (box), whiskers extending to 1.5 × the interquartile range, and individual participant values (points). The sex distribution is presented as proportions. (A) Log-transformed plasma p217tau concentrations. (B) Plasma p217tau/Aβ42 ratio. (C) Plasma Aβ42/Aβ40 ratio. (D) Plasma GFAP. (E) Plasma NfL. (F) Age. (G) Education level. (H) APOE ε4 allele count. (I) Sex distribution. Participants classified as Robust High Risk and Discordant High Risk generally exhibited higher p217tau, p217tau/Aβ42 ratio, GFAP concentrations, and APOE ε4 burden than the Robust Low Risk and Intermediate groups, whereas Aβ42/Aβ40, NfL, age, education, and sex demonstrated comparatively smaller differences across groups. These biomarker profiles provide independent biological support for the consensus-risk classification and are consistent with enrichment of established Alzheimer’s disease- related biomarkers among participants assigned to the higher-risk groups.

The strongest biological differences were observed for p217tau, p217tau/Aβ42, GFAP, and APOE ε4 burden. Participants in the Robust High-Risk and Discordant High-Risk groups exhibited profiles consistent with substantially higher underlying concentrations than participants in the Robust Low Risk and Intermediate groups (Figure 6A). Similarly, the p217tau/Aβ42 ratio increased progressively across the risk spectrum and showed particularly strong separation between low-risk and high-risk participants (Figure 6B). GFAP concentrations were also markedly elevated in the high-risk groups (Figure 6D), consistent with greater astroglial activation or injury-associated biology. APOE ε4 burden increased across the risk spectrum and was especially enriched among Robust High-Risk participants (Figure 6H), supporting the biological plausibility of the consensus-risk classifications.

In contrast, Aβ42/Aβ40 demonstrated only modest separation across groups (Figure 6C), while NfL distributions showed considerable overlap despite a tendency toward higher values in the high-risk groups (Figure 6E). Age and education exhibited relatively small differences across groups (Figure 6F-G), indicating that the consensus- risk framework was not primarily driven by demographic characteristics. Sex distributions varied among groups, although interpretation is limited by the small size of the Robust High-Risk subgroup (Figure 6I).

These findings indicate that consensus-risk classification captures biologically meaningful variation in AD-related plasma biomarkers rather than simply reflecting demographic differences. The enrichment of p217tau, p217tau/Aβ42, GFAP, and APOE ε4 burden among high-risk participants provides independent biological support for the validity of the consensus-risk framework and helps explain why the highest-ranked participants identified by the framework were enriched for established markers of AD-related pathology. These observations are also consistent with the biomarker contribution analyses presented in Figure S5, which identified -related measures, APOE ε4 burden, and GFAP-associated biology as major correlates of consensus risk and framework disagreement.

### Biomarker Contributions to Consensus Risk and Framework Disagreement

To better understand which biological variables were most strongly associated with framework-specific predictions, consensus AD-risk, and prediction uncertainty, we performed model-agnostic permutation importance analyses using surrogate random- forest models trained to reconstruct the outputs of the external machine-learning frameworks (Supplementary Figure S5, Supplementary Tables S7 and S8).

The surrogate models reproduced framework-generated outputs with high fidelity (Table S7). Five-fold cross-validated R² values ranged from 0.82 to 0.97 for framework-specific predictions and reached 0.93 for consensus AD-risk. Mean absolute errors were uniformly low, supporting the ability of the surrogate models to reconstruct framework outputs for subsequent permutation-importance analyses. These results indicate that the selected biomarker and covariate set captured most of the information contained within the framework-generated predictions and supported the validity of the subsequent contribution analyses.

The biological correlates of framework-specific predictions differed substantially.

Complete permutation-importance rankings for all framework outputs are provided in Supplementary Table S8. The surrogate models indicated that A4-derived amyloid probability and predicted centiloid outputs were most strongly associated with p217tau, whereas the ADNI-AlzPath framework outputs were most strongly associated with the p217tau/Aβ42 ratio. In contrast, outputs from the ADNI-Jan framework demonstrated stronger associations with APOE ε4 burden and sex than with p217tau-related measures. These findings suggest that the frameworks emphasize partially distinct biological signals despite being applied to the same participant cohort.

Consensus AD-risk was most strongly associated with p217tau, APOE ε4 burden, and the p217tau/Aβ42 ratio. Prediction uncertainty and rank instability showed broader contributions from APOE ε4 burden, p217tau, p217tau/Aβ42, sex, GFAP, and NfL. APOE ε4 burden ranked among the strongest contributors to both prediction uncertainty and rank instability (Figure S5; Supplementary Table S8). Because these analyses were based on surrogate models trained to reconstruct framework outputs rather than the original machine-learning models, they should be interpreted as exploratory.

## DISCUSSION

In this study, we applied three independently trained machine-learning (ML) frameworks to plasma biomarker measurements generated using the 5ADCSI platform in the Louisville Twins Study (LTS) cohort and evaluated consensus AD-risk across the resulting framework predictions. Four principal findings emerged. First, 5ADCSI-derived biomarker measurements generated biologically coherent amyloid-related predictions across all three reference frameworks. Second, substantial variability existed between frameworks, indicating that both training cohort composition and measurement methodology materially influences downstream ML predictions. Third, despite this variability, a consensus-risk approach identified a stable subgroup of participants who were consistently classified as high risk across independent models while simultaneously quantifying uncertainty for participants whose classification depended strongly on the reference framework. Finally, biological validation and biomarker contribution analyses demonstrated that the consensus-risk framework is anchored in established AD-related biological markers, particularly p217tau, p217tau/Aβ42, APOE ε4 burden, and GFAP. Together, these findings suggest that consensus modeling provides a useful translational strategy for evaluating emerging biomarker platforms and identifying individuals with robust AD-related biological risk signatures. Because the evaluation cohort consisted primarily of middle-aged community-dwelling adults rather than individuals recruited for AD-focused clinical studies, the present findings should be interpreted primarily as evidence of biological transportability across cohorts rather than validation of absolute disease-risk prediction.

### Biological Coherence Is Preserved Across Frameworks

The most fundamental finding of the study is that all three frameworks generated internally coherent predictions despite being trained under different biological and analytical conditions. In the context of this study, biological coherence refers to the extent to which independently trained models recognize the same underlying Alzheimer’s disease-related biological patterns from the 5ADCSI plasma biomarker measurements, even if they differ in their exact numerical predictions.

Within each framework, predicted amyloid positivity probability and predicted centiloid burden remained positively associated, indicating that the models consistently mapped plasma biomarker measurements onto a biologically plausible amyloid-related disease axis. This observation is important because the LTS cohort did not have contemporaneous amyloid PET imaging or clinical outcome data available for direct validation. Consequently, the study was designed to evaluate biological coherence and translational consistency rather than conventional predictive accuracy.

The positive relationships between amyloid probability and centiloid burden suggest that 5ADCSI-derived biomarker measurements contain information that is recognized by independently trained ML models as reflecting underlying AD-related biology. This finding extends prior studies demonstrating the strong biological relevance of plasma p217tau, Aβ42/Aβ40, GFAP, and NfL in predicting amyloid pathology and disease progression^4, 6, 43–45^. While the present study cannot determine whether any specific prediction is correct in the absence of ground-truth pathology, the consistency of these internal relationships supports the conclusion that the 5ADCSI platform captures biologically meaningful signal rather than random variation.

### Framework Agreement Reveals the Impact of Assay Methodology on Blood-Based AD Risk Prediction

Notably, the present study found that framework agreement was strongly dependent on the assay used during model development. Although all three frameworks were applied to identical LTS biomarker measurements, agreement varied substantially across framework pairs.

Multiple studies have reported that p217tau measurements vary according to assay platform, antibody configuration, and analytical methodology^9, 46–48^. The strongest concordance was observed between the A4-MSD and ADNI-AlzPath frameworks. This result was somewhat unexpected because these models differed with respect to both training cohort and assay platform. As a reminder, the A4 framework was developed using p217tau quantified on the Lilly MSD platform, whereas the ADNI-AlzPath framework was developed using p217tau measured on the Quanterix Simoa platform with the AlzPath antibody. Despite these differences, both frameworks frequently identified the same participants as high risk, produced moderately similar continuous prediction outputs, and demonstrated substantial overlap among the highest-risk individuals. This observation suggests that robust AD-related biological information can be recovered across distinct assay technologies when the underlying biological signal is strong.

In contrast, agreement involving the ADNI-Jan framework was consistently weaker. Because the ADNI-AlzPath and ADNI-Jan frameworks were both trained within the same cohort and on the same Quanterix Simoa platform, the principal distinction between these models is the antibody used for quantification. The weaker agreement observed between these frameworks therefore suggests that antibody-specific differences may influence the biological relationships learned by downstream ML models. Importantly, these differences affected not only continuous prediction values but also which participants were ultimately prioritized as highest risk. Surprisingly, the strongest overlap was observed between A4-MSD and ADNI-AlzPath frameworks despite differences in both training cohort and assay platform, whereas overlap between the two ADNI-derived frameworks was substantially weaker. These findings indicate that framework agreement cannot be explained solely by cohort composition or assay platform and suggest that antibody-specific measurement characteristics may exert a meaningful influence on downstream prediction behavior.

### Framework Agreement and Biological Signal Are Not the Same Concept

A central observation of this study is that biological coherence and cross-framework agreement are not equivalent. Although all three frameworks produced plausible amyloid-related predictions, agreement between frameworks varied substantially. In some comparisons, participant-level correlations approached zero despite the fact that each framework independently generated coherent amyloid-related outputs. This distinction is important because biomarker-platform evaluation is often approached primarily through direct assay correlation or prediction agreement.

However, calibration, transportability, and biological validity represent distinct properties of predictive models^49–52^. The present findings suggest that agreement metrics alone may not fully capture the biological value of a novel biomarker platform. A measurement system may preserve meaningful disease-related information even when predictions generated by different external frameworks differ quantitatively. Accordingly, the critical observation is not perfect agreement among models, but rather their ability to detect a common underlying AD-related biological signal despite differences in prediction scale and participant ranking.

### Consensus Risk Provides More Information Than Any Individual Framework

The primary methodological contribution of this study is the introduction of a consensus-risk framework that integrates predictions from multiple independent biological reference models while explicitly quantifying prediction uncertainty. Rather than viewing disagreement among independently trained frameworks as a limitation, the consensus approach treats that disagreement as an additional source of biological information. The framework therefore estimates both participant-level AD-related biological risk and the degree of confidence in that estimate across independent reference models.

Consensus and ensemble-learning approaches have long been used to improve the robustness and generalizability of predictive models by integrating complementary information across multiple independently developed models^53, 54^. More recently, disagreement among models has been recognized as a useful measure of predictive uncertainty, particularly when models are developed under different assumptions or training conditions^53, 55, 56^. Although our framework differs from traditional ensemble-learning methods because it combines externally developed models trained in different cohorts and using different p217tau assays, it is based on the same general principle that integrating complementary sources of information can provide a more robust characterization than reliance on any single model alone.

The consensus framework identified two distinct forms of elevated AD-related biological risk. Robust High Risk participants exhibited high consensus-risk scores together with low prediction uncertainty and low rank instability, indicating that their classification was largely independent of differences in training cohort, assay methodology, and learned biomarker relationships. In contrast, Discordant High Risk participants demonstrated similarly elevated consensus-risk scores but substantially greater prediction uncertainty and rank instability, indicating that their classification remained sensitive to differences among the external reference frameworks. These findings suggest that elevated biological risk and confidence in that risk estimate represent related but distinct dimensions of participant characterization.

An important translational implication of this framework is that disagreement among independently trained models should not necessarily be viewed as model failure. Instead, participants classified as Discordant High Risk may represent biologically informative cases in which AD-related biomarker profiles occupy transitional, heterogeneous, or model-sensitive regions of the disease continuum. These participants exhibit elevated consensus-risk estimates but greater uncertainty because different biological reference frameworks interpret their biomarker profiles differently.

Such individuals may therefore represent particularly valuable targets for longitudinal follow-up, multimodal biomarker assessment, or future pathological validation, as they may provide insight into biological processes that are not fully captured by any single predictive model.

Rather than replacing individual framework predictions, the combination of consensus risk and prediction uncertainty provides a richer characterization of participant-level AD-related biological risk by explicitly identifying both the magnitude and the stability of the estimated risk. This information is not available from any individual framework alone and may prove valuable when evaluating emerging blood- based biomarker platforms in the absence of direct pathological validation.

### Biological Validation Supports the Consensus-Risk Framework

A major strength of the consensus-risk approach is that the highest-ranked participants demonstrated enrichment for established AD-related biological markers. Participants prioritized by the framework exhibited greater APOE ε4 burden together with markedly elevated p217tau, p217tau/Aβ42, and GFAP, all established markers of AD-related pathology and disease progression^2, 57^. In contrast, age differed little between groups, indicating that participant prioritization was not simply driven by demographic factors.

The biomarker and covariate analyses extended these observations across the entire cohort. Robust High Risk and Discordant High Risk participants demonstrated elevated, p217tau, p217tau/Aβ42, GFAP, and APOE ε4 burden relative to lower-risk groups, whereas Aβ42/Aβ40 and NfL showed comparatively weaker separation.

Together, these findings support the biological validity of the consensus-risk classifications.

### Biomarker Contribution Analyses Suggest Potential Sources of Framework Disagreement

The exploratory biomarker contribution analyses provide insight into why independently trained frameworks produced different risk estimates despite being applied to identical LTS biomarker measurements. Although these analyses were based on surrogate models trained to reconstruct framework outputs--using global surrogate models, a well-established model-agnostic approach for interpreting otherwise inaccessible prediction models^41, 58^--rather than on the original machine-learning models themselves, they revealed notable differences in the biological signals associated with each framework. Although these analyses were based on surrogate rather than the original machine-learning models, global surrogate modeling provides an established model-agnostic approach for interpreting otherwise inaccessible prediction models^41, 58^.

The A4-derived framework outputs were most strongly associated with p217tau, whereas the ADNI-AlzPath framework outputs were most strongly associated with the p217tau/Aβ42 ratio. In contrast, the ADNI-Jan framework outputs showed stronger associations with APOE ε4 burden and sex. These observations suggest that the frameworks are not simply scaled versions of one another but instead appear to emphasize partially distinct biological features when generating risk estimates.

Taken together, these biomarker contribution patterns provide a plausible explanation for why independently trained frameworks frequently disagreed despite being applied to identical biomarker measurements. Rather than emphasizing identical biological features, each framework appears to weigh complementary aspects of the underlying plasma biomarker profile. Consequently, participants with complex or heterogeneous biological signatures may receive similar overall consensus-risk estimates while differing in the degree to which individual frameworks prioritize them.

In addition to APOE ε4 burden, p217tau-related measures, GFAP, and NfL were associated with prediction uncertainty and rank instability, suggesting that disagreement among frameworks is influenced by multiple biological processes rather than a single biomarker. This broader pattern is consistent with the multidimensional nature of Alzheimer’s disease biology and further supports the interpretation that framework disagreement reflects biological heterogeneity rather than random model variability.

Perhaps most importantly, APOE ε4 burden emerged as one of the strongest variables associated with both prediction uncertainty and rank instability. In addition to APOE ε4 burden, p217tau-related measures, GFAP, and NfL were also associated with prediction uncertainty and rank instability, suggesting that disagreement among frameworks is influenced by multiple biological processes rather than a single biomarker. These observations indicate that framework disagreement is associated with biologically meaningful participant characteristics rather than random prediction variability and are consistent with the multidimensional nature of Alzheimer’s disease biology.

## Limitations

Several limitations should be considered when interpreting these findings. First, amyloid PET imaging was not available in the LTS cohort, preventing direct evaluation of framework predictions against an independent biological reference standard.

Consequently, the study evaluated biological coherence and consensus-risk behavior rather than conventional predictive accuracy.

Second, although the consensus framework integrated predictions from three independently trained machine-learning models developed in two external cohorts (ADNI and A4), all participant-level evaluations were performed in a single independent cohort (LTS). Additional studies in independent cohorts with different demographic characteristics, disease prevalence, and biomarker distributions will be necessary to establish the generalizability and robustness of the proposed consensus-risk framework.

Third, differences observed among the external frameworks likely reflect the combined effects of training cohort composition, p217tau assay methodology, preprocessing strategies, model architecture, and learned biomarker relationships, making it difficult to isolate the contribution of any individual factor. Consequently, the present analyses should be interpreted as evaluating the combined influence of these factors rather than the independent effect of cohort characteristics or assay methodology alone.

Fourth, the biomarker contribution analyses were exploratory and were based on surrogate machine-learning models rather than direct feature-importance estimates extracted from the original externally trained frameworks. Although surrogate models provide a well-established model-agnostic approach for interpreting otherwise inaccessible prediction models, the resulting feature contributions should be viewed as approximate representations of framework behavior rather than exact explanations of the original models. Nevertheless, the consistency of the observed biomarker associations with established AD biology supports the biological plausibility of the consensus-risk framework and reinforces the central role of p217tau-related biology in blood-based AD risk assessment.

Fifth, although the present analyses demonstrate that 5ADCSI-derived plasma biomarker measurements preserve biologically meaningful AD-related signal across multiple independent reference frameworks, they do not establish the relative predictive accuracy of either the individual frameworks or the consensus framework in the absence of an independent biological reference standard. Accordingly, the consensus- risk framework should be viewed as a complementary strategy for integrating multiple external biological reference models and quantifying prediction uncertainty rather than as a replacement for direct pathological or imaging-based validation.

Finally, an additional consideration is that the LTS cohort was substantially younger than the populations used to develop the external reference frameworks. The A4 framework was trained primarily in cognitively unimpaired older adults, whereas the ADNI-derived frameworks included older individuals spanning the Alzheimer’s disease continuum. In contrast, the LTS cohort represents a community-based middle-aged population with a lower expected prevalence of AD-related pathology. Consequently, differences in age distribution and disease prevalence may influence the absolute calibration of framework predictions. However, because the objective of the present study was to evaluate biological coherence, framework agreement, and consensus-risk behavior rather than absolute prediction accuracy, these demographic differences are unlikely to alter the primary conclusions regarding preservation of biologically meaningful AD-related signal across independently trained reference frameworks.

Future studies should evaluate the consensus-risk framework in older community-based cohorts with contemporaneous amyloid PET or longitudinal clinical follow-up.

## Conclusions

The present study demonstrates that plasma biomarker measurements generated using the 5ADCSI platform contain biologically meaningful AD-related information that is consistently recognized across multiple independently trained machine-learning frameworks. Although framework-specific predictions differed according to training cohort and p217tau assay methodology, integrating these independent biological reference frameworks through a consensus-risk approach identified a stable subgroup of participants with reproducibly elevated AD-related biological risk while simultaneously quantifying prediction uncertainty arising from framework disagreement.

These findings suggest that consensus-risk modeling provides a practical strategy for interpreting emerging blood-based biomarker platforms by combining complementary information from multiple external reference models while explicitly characterizing the stability of participant-level risk estimates. More broadly, the combination of consensus risk, prediction uncertainty, and rank stability offers a biologically informed framework for evaluating novel plasma biomarker technologies and prioritizing individuals for future Alzheimer’s disease research, longitudinal follow- up, and eventual clinical translation when direct pathological validation is unavailable.

## Acknowledgments

Data used in the preparation of this article were obtained from the Alzheimer’s Disease Neuroimaging Initiative (ADNI) database (adni.loni.usc.edu). ADNI is funded by the National Institute on Aging, the National Institute of Biomedical Imaging and Bioengineering, and through contributions from industry partners. Data was also obtained from the Anti-Amyloid Treatment in Asymptomatic Alzheimer’s Disease (A4) Study. The A4 Study is funded by the National Institute on Aging, Eli Lilly and Company, the Alzheimer’s Association, and additional philanthropic contributors.

## Ethical considerations

All study procedures were conducted in accordance with the Declaration of Helsinki and were approved by the Institutional Review Board at the University of Louisville (19.0989) and the University of Southern California (UP-20-00029). All plasma samples were deidentified prior to analyses. Written informed consent was obtained from all participants or their legally authorized representatives prior to participation.

## Author’s Contributions

EZ conceived and designed the study, conducted the investigation and formal analyses, quantified the plasma biomarkers for the LTS cohort, developed the machine learning models, supervised the project, generated the figures and tables, interpreted the data, drafted and wrote the original manuscript. All authors read and approved of the final manuscript.

## Data availability

The data supporting the findings of this study are available from corresponding author upon reasonable request from educational institutions only.

## Supplemental material

Supplemental material for this article is available online.

## Supplementary Figures & Legends

**Figure S1.**
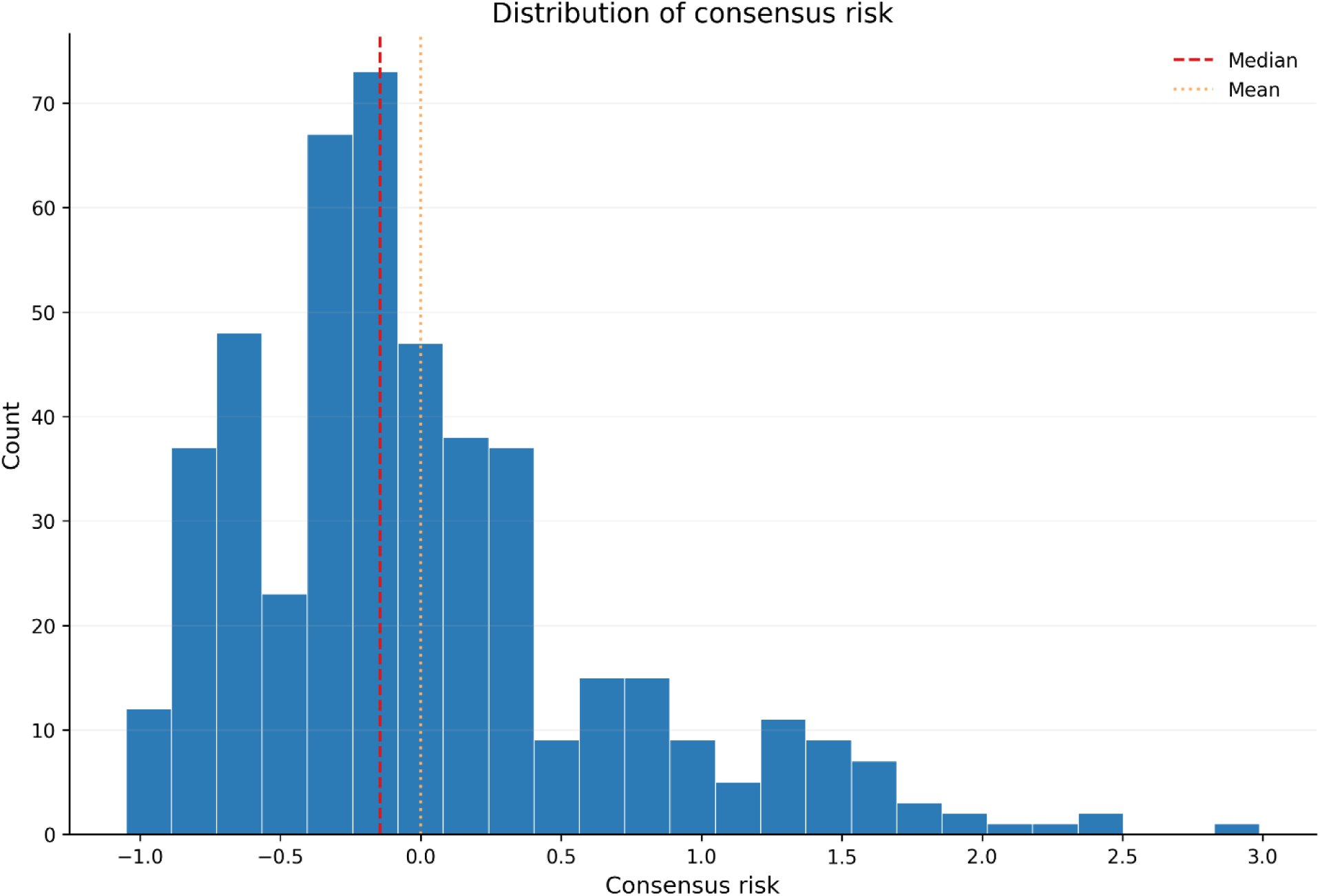
Distribution of Consensus AD-Risk Scores. Histogram showing the distribution of participant-level consensus AD-risk scores across the Longitudinal Twins Study (LTS) cohort. Consensus AD-risk was calculated as the average of consensus amyloid risk and consensus centiloid burden after framework-specific standardization of predictions generated by the A4-MSD, ADNI-AlzPath, and ADNI-Jan machine-learning models. The red dashed line indicates the median consensus-risk score and the orange dotted line indicates the mean consensus-risk score. The distribution is positively skewed, with most participants concentrated around low-to-intermediate consensus-risk values and a relatively small subset of participants occupying the high-risk tail of the distribution. This pattern is consistent with the identification of a limited number of participants with markedly elevated consensus AD- risk provides context for the consensus-risk group classification presented in Figure 2.

**Figure S2.**
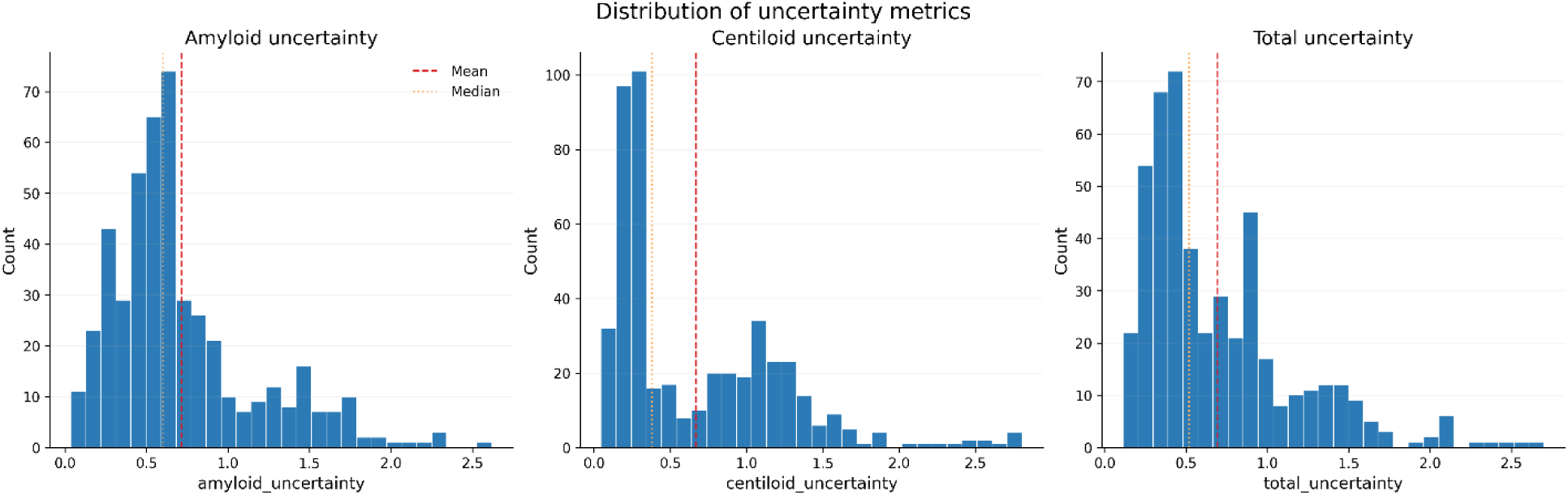
Distribution of Prediction Uncertainty Metrics. Histograms showing the distributions of amyloid uncertainty, predicted centiloid uncertainty, and total uncertainty across the LTS cohort. Uncertainty was calculated as the standard deviation of predictions generated by the three external machine-learning frameworks. These distributions provide context for the uncertainty analyses presented in Figure 4.

**Figure S3.**
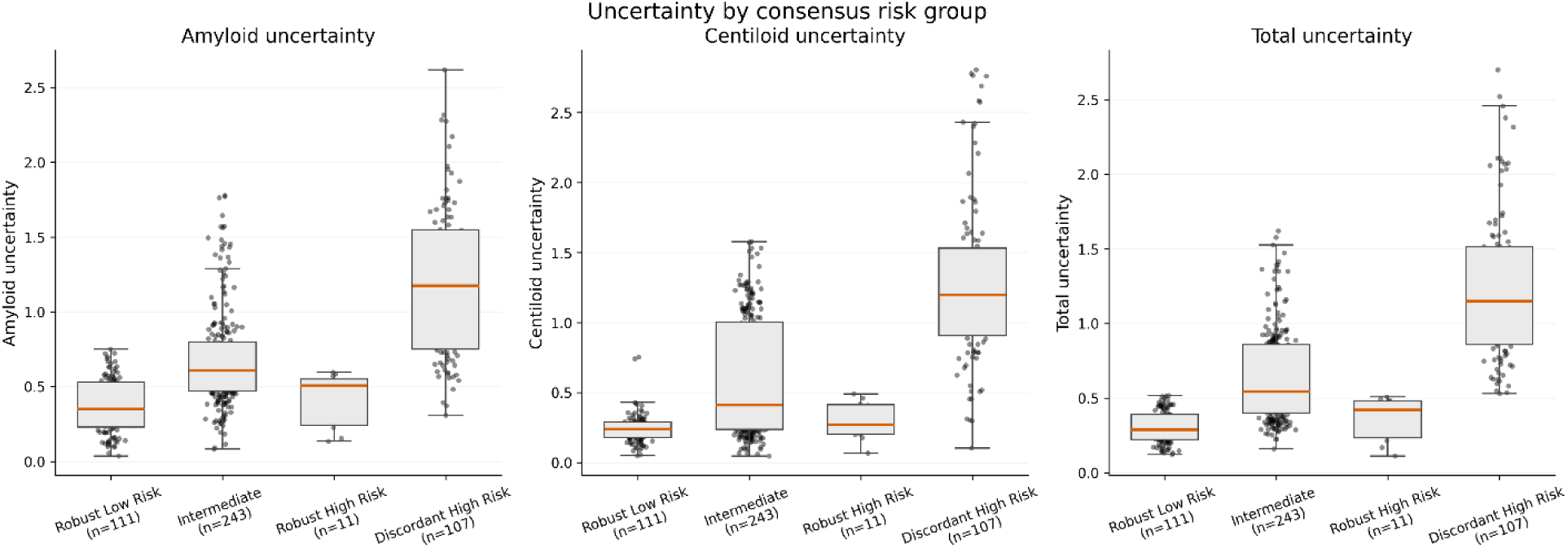
Prediction Total Uncertainty Across Consensus Risk Groups. Boxplots of amyloid uncertainty, centiloid uncertainty, and total uncertainty are stratified by consensus-risk group. Discordant High Risk participants exhibit substantially greater framework disagreement than Robust High Risk or Robust Low Risk participants. These analyses complement the consensus-risk classifications shown in Figure 2.

**Figure S4.**
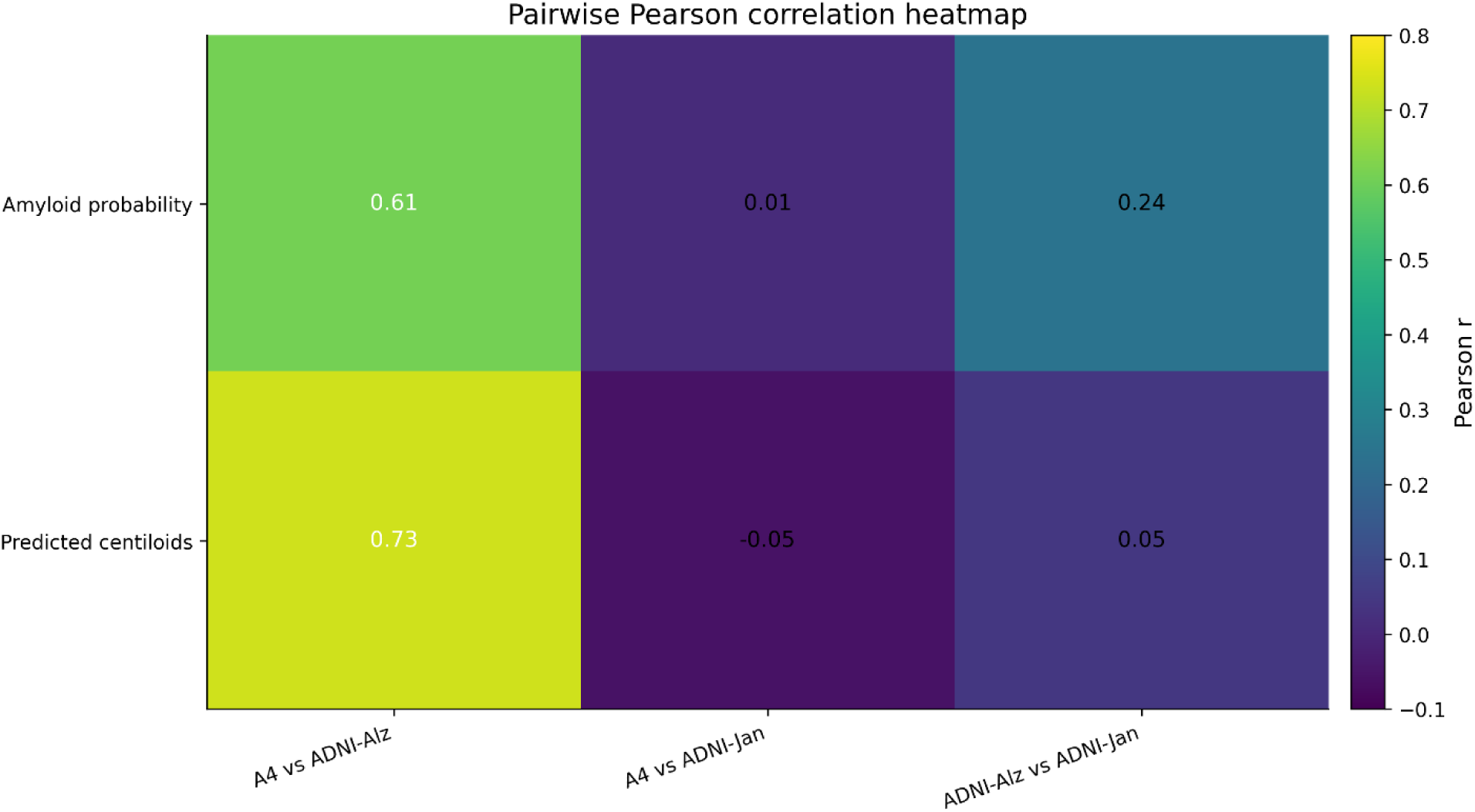
Pairwise Pearson Correlation Heatmap. Pairwise Pearson correlations comparing continuous prediction outputs generated for identical LTS participants by the A4-MSD, ADNI-AlzPath, and ADNI-Jan frameworks. Strongest agreement was observed between the A4-MSD and ADNI-AlzPath frameworks. These analyses correspond to the framework agreement metrics summarized in Supplementary Table S4.

**Figure S5.**
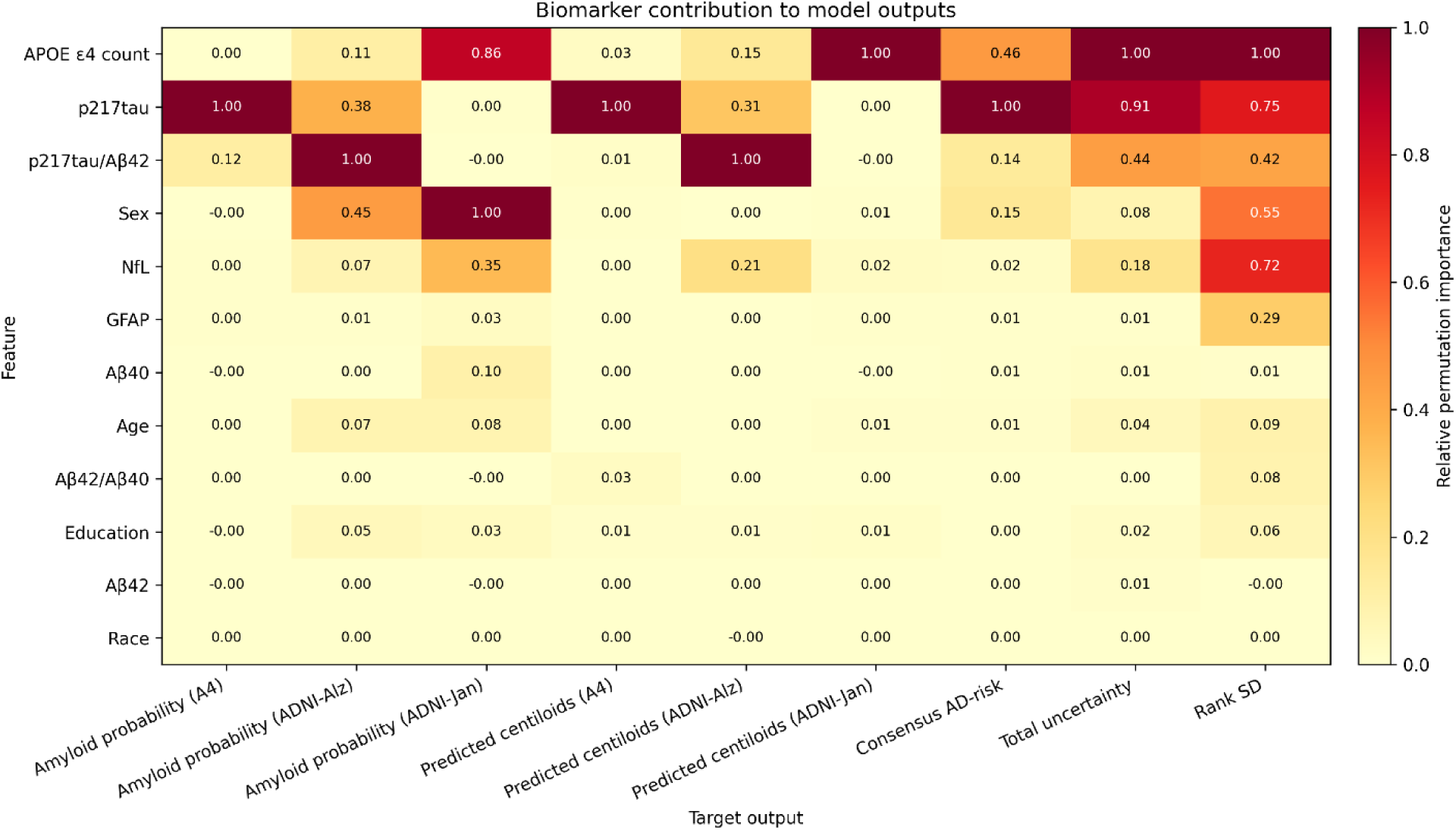
Biomarker Contributions to Framework-Specific Predictions, Consensus Risk, and Prediction Uncertainty. Heatmap summarizing relative biomarker and covariate contributions to framework-specific prediction outputs, consensus AD-risk, prediction uncertainty, and rank instability. Values are normalized independently within each column and therefore should be interpreted only within each prediction target. Contributions were estimated using model-agnostic permutation importance analyses applied to surrogate random- forest models trained to reconstruct prediction outputs generated by the external machine-learning frameworks. Columns represent prediction targets, including amyloid probability predictions from the A4, ADNI-AlzPath, and ADNI-Jan frameworks; predicted centiloid burden from each framework; composite consensus AD-risk; total prediction uncertainty; and rank instability quantified as rank standard deviation (Rank SD). Rows represent demographic variables, APOE ε4 burden, and plasma biomarker measurements. Values indicate relative permutation importance normalized within each prediction target, such that the most influential feature for a given target receives a value of 1.0. For the A4 framework, p217tau was the dominant contributor to both amyloid probability and predicted centiloid outputs. In contrast, the ADNI-AlzPath framework relied most strongly on the p217tau/Aβ42 ratio, whereas the ADNI-Jan framework was driven primarily by APOE ε4 burden and sex. Consensus AD-risk was predominantly associated with p217tau, APOE ε4 burden, and p217tau/Aβ42. Prediction uncertainty and rank instability were associated with a broader set of variables, including APOE ε4 burden, p217tau, p217tau/Aβ42, sex, GFAP, and NfL. These exploratory analyses complement the biological interpretation presented in the Results and Discussion.

## Supplementary Tables

**Table S1.** Descriptive summary of framework-specific prediction outputs prior to consensus-risk standardization. Summary statistics for framework-specific predictions generated from the same LTS participant biomarker data using three external machine-learning reference frameworks: A4-MSD, ADNI-AlzPath, and ADNI-Jan. For each framework, summary statistics are provided for predicted amyloid positivity probability and predicted centiloid burden. Values are reported as mean ± SD and median (IQR).

| Framework | Metric | N | Mean ± SD | Median (IQR) | Min | Max |
| --- | --- | --- | --- | --- | --- | --- |
| A4-MSD | Amyloid probability | 472 | 0.26 ± 0.42 | 0.00 (0.00–0.62) | 0.00 | 1.00 |
| A4-MSD | Predicted centiloids | 472 | 49.83 ± 25.72 | 40.14 (32.15–56.27) | 14.90 | 139.77 |
| ADNI-AlzPath | Amyloid probability | 472 | 0.32 ± 0.08 | 0.30 (0.26–0.36) | 0.17 | 0.75 |
| ADNI-AlzPath | Predicted centiloids | 472 | 14.24 ± 16.00 | 10.68 (7.69–13.38) | -8.57 | 87.22 |
| ADNI-Jan | Amyloid probability | 472 | 0.24 ± 0.07 | 0.24 (0.18–0.27) | 0.10 | 0.47 |
| ADNI-Jan | Predicted centiloids | 472 | 15.44 ± 15.03 | 8.99 (5.28–31.83) | -6.74 | 50.35 |

**Table S2.** Framework-Specific Prediction Uncertainty Across Consensus Risk Groups. Values are presented as Mean ± SD and Median (IQR). N (%) indicates the number and percentage of participants in each consensus-risk group. Summary of amyloid uncertainty, centiloid uncertainty, total uncertainty, and rank instability across the four consensus AD-risk groups. Amyloid uncertainty and centiloid uncertainty were calculated as the standard deviation of framework-specific predictions. Total uncertainty was calculated as the average of these two measures. Rank instability was calculated as the standard deviation of participant ranking across frameworks. Robust Low Risk and Robust High Risk participants exhibit low uncertainty, whereas Discordant High Risk participants demonstrate substantially greater disagreement among frameworks. These analyses provide the numerical basis for the uncertainty distributions shown in Figure 2E and Supplementary Figure S3.

| Risk Group | N (%) | Amyloid |  | Centiloid |  | Total |  | Rank SD |  |
| --- | --- | --- | --- | --- | --- | --- | --- | --- | --- |
| | | Mean ± SD | Median (IQR) | Mean ± SD | Median (IQR) | Mean ± SD | Median (IQR) | Mean ± SD | Median (IQR) |
| Robust Low Risk | 111 (23.5) | 0.37 ± 0.18 | 0.35 (0.23–0.53) | 0.24 ± 0.10 | 0.24 (0.18–0.29) | 0.31 ± 0.11 | 0.29 (0.22–0.39) | 74 ± 30 | 69 (51–100) |
| Intermediate | 243 (51.5) | 0.68 ± 0.33 | 0.61 (0.47–0.80) | 0.60 ± 0.43 | 0.41 (0.24–1.00) | 0.64 ± 0.31 | 0.54 (0.40–0.86) | 107 ± 34 | 103 (80–129) |
| Robust High Risk | 11 (2.3) | 0.42 ± 0.18 | 0.51 (0.24–0.55) | 0.30 ± 0.14 | 0.27 (0.20–0.41) | 0.36 ± 0.15 | 0.42 (0.23–0.48) | 25 ± 8 | 26 (21–29) |
| Discordant High Risk | 107 (22.7) | 1.18 ± 0.50 | 1.17 (0.75–1.55) | 1.29 ± 0.59 | 1.20 (0.91–1.53) | 1.24 ± 0.50 | 1.15 (0.86–1.51) | 125 ± 58 | 136 (72–172) |

Values are presented as Mean ± SD and Median (IQR). N (%) indicates the number and percentage of participants in each consensus-risk group.

**Table S3.** Full Consensus-Risk Ranking of LTS Participants. Participant-level ranking generated by the consensus-risk framework. For each participant, the table reports consensus AD-risk score, consensus amyloid risk, consensus centiloid burden, prediction uncertainty, rank instability, and consensus-risk group assignment. Consensus-risk scores were generated by integrating framework-specific predictions from the A4-MSD, ADNI-AlzPath, and ADNI-Jan machine-learning models. Participants were ranked from highest to lowest consensus-risk score. This table represents the complete output of the consensus-risk framework and provides the participant-level data underlying the ranking, uncertainty, and risk-group analyses presented throughout the manuscript.

**Table S4.**
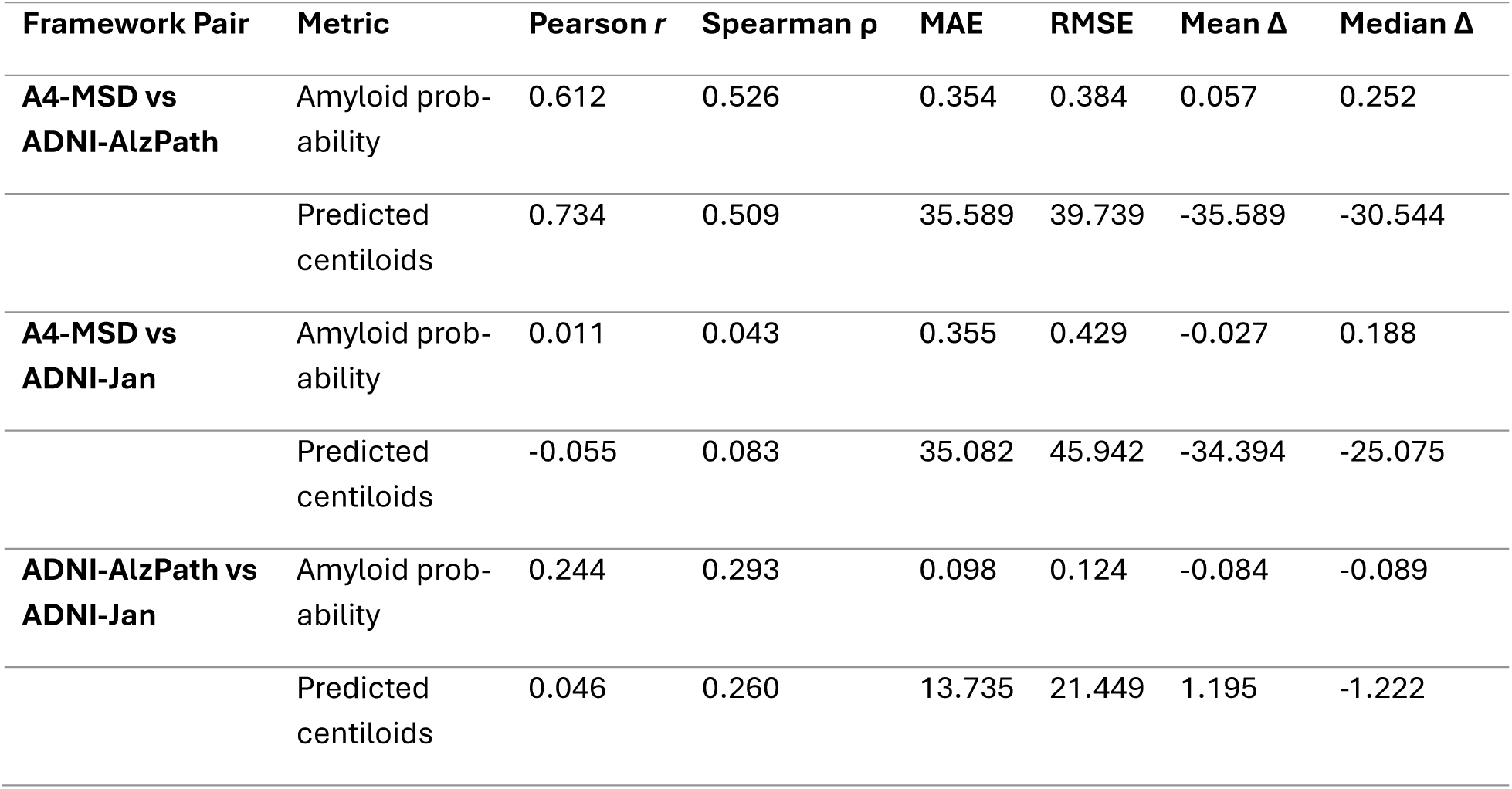
Pairwise Agreement of Framework-Specific Predictions. Pairwise comparison of continuous prediction outputs generated for the same LTS participants by the A4-MSD, ADNI-AlzPath, and ADNI-Jan machine-learning frameworks. Agreement was evaluated separately for predicted amyloid positivity probability and predicted centiloid burden. Pearson correlations quantify linear agreement, Spearman correlations quantify rank-order agreement, mean absolute error (MAE) quantifies average prediction differences, and root mean squared error (RMSE) quantifies overall prediction divergence. Importantly, these comparisons evaluate agreement among framework-specific interpretations of identical LTS biomarker measurements rather than comparisons between the original ADNI and A4 cohorts. Agreement is strongest between the two AlzPath-based frameworks and substantially weaker in comparisons involving the Jan-based framework, suggesting that p217tau assay platform influences downstream machine-learning predictions.

**Table S5.** Agreement of Framework–Derived Centiloid Categories. Agreement between categorical centiloid classifications assigned to LTS participants by different machine–learning frameworks. Continuous predicted centiloid values were converted to categorical centiloid groups using predefined thresholds. Agreement was quantified using both raw agreement percentage and Cohen’s κ statistic. Although some framework comparisons demonstrate moderate raw agreement, κ values remain close to zero, indicating that much of the apparent agreement is attributable to category prevalence rather than true participant–level concordance. These findings support the use of continuous prediction measures rather than threshold-derived categories in the consensus-risk framework.

| Pair | N valid | Agreement | Kappa |
| --- | --- | --- | --- |
| A4-MSD vs ADNI-AlzPath | 472 | 0.0296 | -0.0523 |
| A4-MSD vs ADNI-Jan | 472 | 0.1080 | -0.0200 |
| ADNI-AlzPath vs ADNI-Jan | 472 | 0.625 | 0.0330 |

**Table S6.** Overlap of Highest-Risk Participants Across Frameworks. Top-k overlap analysis evaluating whether different machine-learning frameworks identify the same LTS participants as highest risk. Participants were ranked according to predicted amyloid positivity probability and predicted centiloid burden. Pairwise overlap was calculated for the top 10, 25, 50, and 100 participants identified by each framework. Jaccard similarity and overlap percentage were calculated to quantify concordance. Overlap is substantially greater between the two AlzPath-based frameworks than in comparisons involving the Jan-based framework. These results indicate that participant prioritization is sensitive to the p217tau assay platform used during model development and support the rationale for consensus-risk modeling.

| Pair | Metric | k | Overlap n | Jaccard | Overlap % of k |
| --- | --- | --- | --- | --- | --- |
| A4_MSD vs ADNI-AlzPath | Amyloid probability | 10 | 6 | 0.42857<br>14 | 0.6 |
| A4_MSD vs ADNI-AlzPath | Amyloid probability | 25 | 20 | 0.66666<br>67 | 0.8 |
| A4_MSD vs ADNI-AlzPath | Amyloid probability | 50 | 33 | 0.49253<br>73 | 0.66 |
| A4_MSD vs ADNI-AlzPath | Amyloid probability | 10<br>0 | 63 | 0.45985<br>4 | 0.63 |
| A4_MSD vs ADNI-AlzPath | Predicted centiloids | 10 | 6 | 0.42857<br>14 | 0.6 |
| A4_MSD vs ADNI-AlzPath | Predicted centiloids | 25 | 12 | 0.31578<br>95 | 0.48 |
| A4_MSD vs ADNI-AlzPath | Predicted centiloids | 50 | 33 | 0.49253<br>73 | 0.66 |
| A4_MSD vs ADNI-AlzPath | Predicted centiloids | 10<br>0 | 83 | 0.70940<br>17 | 0.83 |
| A4_MSD vs ADNI-Jan | Amyloid probability | 10 | 0 | 0 | 0 |
| A4_MSD vs ADNI-Jan | Amyloid probability | 25 | 1 | 0.02040<br>82 | 0.04 |
| A4_MSD vs ADNI-Jan | Amyloid probability | 50 | 7 | 0.07526<br>88 | 0.14 |
| A4_MSD vs ADNI-Jan | Amyloid probability | 10<br>0 | 21 | 0.11731<br>84 | 0.21 |
| A4_MSD vs ADNI-Jan | Predicted centiloids | 10 | 0 | 0 | 0 |
| A4_MSD vs ADNI-Jan | Predicted centiloids | 25 | 1 | 0.02040<br>82 | 0.04 |
| A4_MSD vs ADNI-Jan | Predicted centiloids | 50 | 2 | 0.02040<br>82 | 0.04 |
| A4_MSD vs ADNI-Jan | Predicted centiloids | 10<br>0 | 14 | 0.07526<br>88 | 0.14 |
| ADNI-AlzPath vs ADNI-Jan | Amyloid probability | 10 | 0 | 0 | 0 |
| ADNI-AlzPath vs ADNI-Jan | Amyloid probability | 25 | 2 | 0.04166<br>67 | 0.08 |
| ADNI-AlzPath vs ADNI-Jan | Amyloid probability | 50 | 16 | 0.19047<br>62 | 0.32 |
| ADNI-AlzPath vs ADNI-Jan | Amyloid probability | 10<br>0 | 29 | 0.16959<br>06 | 0.29 |
| ADNI-AlzPath vs ADNI-Jan | Predicted centiloids | 10 | 1 | 0.05263<br>16 | 0.1 |
| ADNI-AlzPath vs<br>ADNI-Jan | Predicted<br>centiloids | 25 | 2 | 0.04166<br>67 | 0.08 |
| ADNI-AlzPath vs<br>ADNI-Jan | Predicted<br>centiloids | 50 | 4 | 0.04166<br>67 | 0.08 |
| ADNI-AlzPath vs<br>ADNI-Jan | Predicted<br>centiloids | 10<br>0 | 22 | 0.12359<br>55 | 0.22 |

**Table S7.**
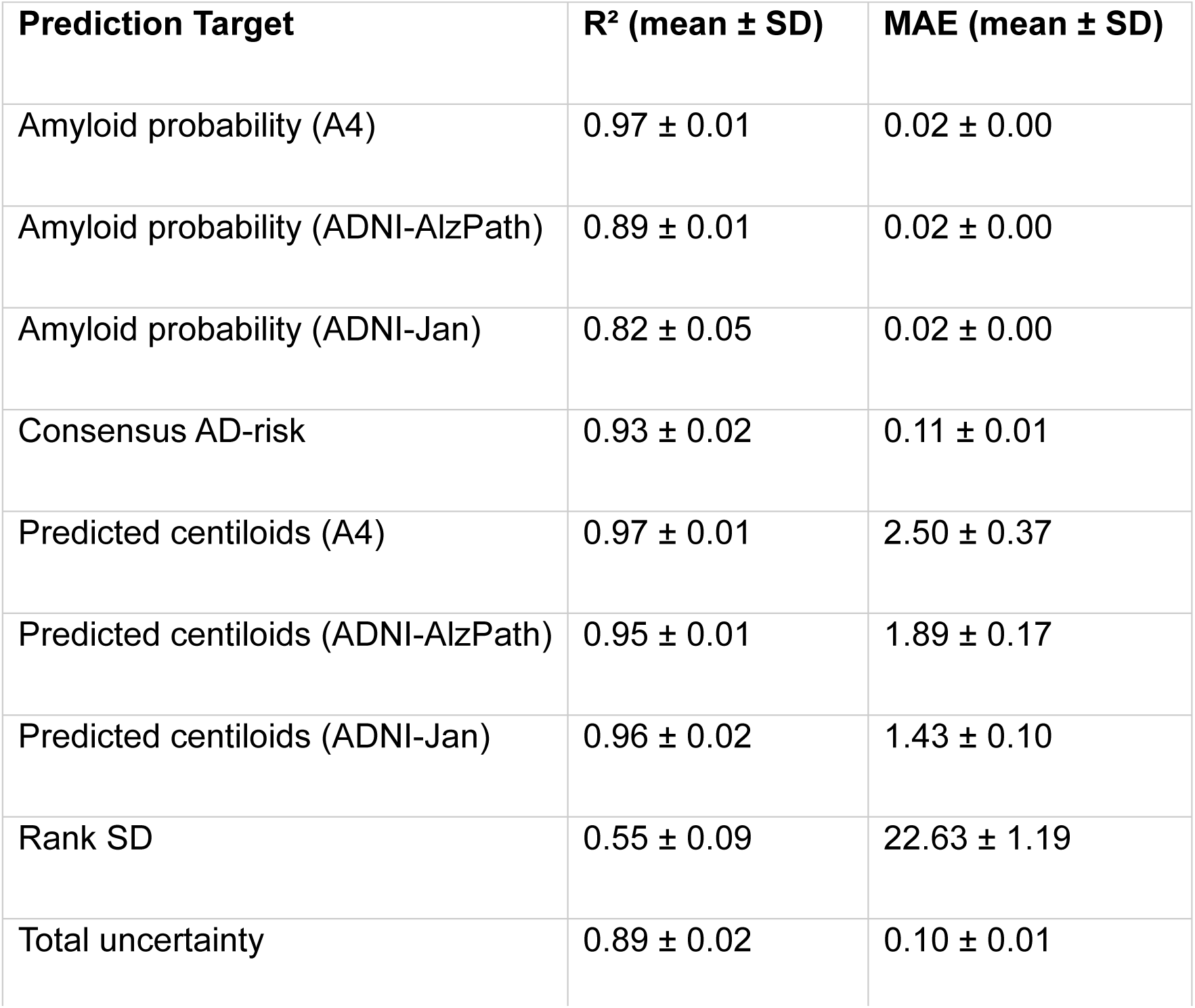
Surrogate Model Fidelity. Performance of surrogate random-forest models used for biomarker contribution analyses. Five-fold cross-validation was performed for all surrogate models (N = 472 participants). For each prediction target, a surrogate model was trained using demographic variables, APOE ε4 burden, and plasma biomarkers to reconstruct framework-generated prediction outputs. Five-fold cross-validation was used to estimate model performance. Fidelity was quantified using the coefficient of determination (R^2^) and mean absolute error (MAE). Columns are defined as follows:

| Prediction Target | R<sup>2</sup> (mean ± SD) | MAE (mean ± SD) |
| --- | --- | --- |
| Amyloid probability (A4) | 0.97 ± 0.01 | 0.02 ± 0.00 |
| Amyloid probability (ADNI-AlzPath) | 0.89 ± 0.01 | 0.02 ± 0.00 |
| Amyloid probability (ADNI-Jan) | 0.82 ± 0.05 | 0.02 ± 0.00 |
| Consensus AD-risk | 0.93 ± 0.02 | 0.11 ± 0.01 |
| Predicted centiloids (A4) | 0.97 ± 0.01 | 2.50 ± 0.37 |
| Predicted centiloids (ADNI-AlzPath) | 0.95 ± 0.01 | 1.89 ± 0.17 |
| Predicted centiloids (ADNI-Jan) | 0.96 ± 0.02 | 1.43 ± 0.10 |
| Rank SD | 0.55 ± 0.09 | 22.63 ± 1.19 |
| Total uncertainty | 0.89 ± 0.02 | 0.10 ± 0.01 |

- R^2^ mean: Mean coefficient of determination across folds.
- R^2^ SD: Standard deviation of R^2^ across folds.
- MAE mean: Mean absolute error across folds.
- MAE SD: Standard deviation of MAE across folds. Surrogate models demonstrated excellent reconstruction of framework-specific prediction outputs. Mean cross-validated R^2^ values ranged from 0.82 to 0.97 for framework-specific amyloid probability and centiloid predictions, 0.93 for consensus AD-risk, and 0.89 for total uncertainty. Rank instability was more difficult to reconstruct (R^2^ = 0.55), indicating that framework disagreement likely reflects more complex interactions among biomarkers and model-specific characteristics.

**Table S8.** Biomarker Contribution Analysis. Permutation-based feature contribution analysis for framework-specific prediction outputs, consensus AD-risk, prediction uncertainty, and rank instability. Because the original machine-learning frameworks were externally trained and not directly available for feature-importance extraction, model-agnostic surrogate random-forest models were trained to reconstruct each prediction output using demographic variables, APOE ε4 burden, and plasma biomarker measurements from the LTS cohort. Separate surrogate models were trained for amyloid probability predictions, predicted centiloid burden, consensus AD-risk, total uncertainty, and rank instability. Feature importance was quantified using permutation importance. For each feature, values were randomly permuted while all other variables were held constant, and the resulting reduction in model performance was measured. Greater reductions in model performance indicate stronger contribution to the target prediction. Relative importance values were normalized within each target output so that the largest contributor received a value of 1.0. Columns are defined as follows:

- Target: Prediction output being reconstructed.
- Target column: Original prediction variable used in the analysis.
- Feature: Variable evaluated for contribution.
- Feature label: Human-readable feature name.
- Importance mean: Mean permutation importance across cross-validation folds.
- Importance SD: Standard deviation of importance estimates across folds.
- Importance relative: Importance normalized to the most influential feature within each target.
- Rank: Relative ranking of feature contribution within each target. The analysis demonstrates substantial differences in the biological variables driving framework-specific predictions. A4-derived predictions were primarily influenced by, whereas ADNI-AlzPath predictions were more strongly associated with the /Aβ42 ratio. The ADNI-Jan framework showed stronger dependence on APOE ε4 burden and sex. Consensus AD-risk was driven predominantly by -related measures and APOE ε4 burden, whereas uncertainty and rank instability reflected contributions from multiple b

